# Applying the Theoretical Domains Framework to the Implementation of Medicaid Coverage for Doula Services: Doulas’ Perceptions of Barriers and Facilitators in the State of Florida

**DOI:** 10.1101/2024.01.06.24300904

**Authors:** Janelle Applequist, Roneé Wilson, Megan Perkins, Victoria Evans, Angela Daniel, Averjill Rookwood, Elizabeth Simmons, Adetola F. Louis-Jacques

## Abstract

As of September 2023, more than half of US states have either expanded Medicaid to cover doula care or are in the process of implementing doula care into Medicaid services. However, little is known about doulas’ experiences with Medicaid. Doula uptake of these services is critical to support low-income clients. We explored Florida doulas’ experiences with and perceptions of Medicaid reimbursement. We conducted seven focus groups and seven in-depth interviews with 31 doulas providing perinatal support. An inductive approach to thematic analysis was used to generate primary findings. A deductive approach was then applied to these themes, drawing on the Theoretical Domains Framework to enhance understanding. Most participants reported wanting to engage with Medicaid to support marginalized communities, but not doing so due to misinformation, low reimbursement rates and a complicated reimbursement structure. Doulas that work with Medicaid patients reflected on their frustrations with insurers and the financial impact of delayed reimbursements. Misinformation spread amongst doula communities about the Medicaid reimbursement process (often based on outdated information) was found to be a significant barrier to implementation. Doulas relied on one another for information, as they reported few resources have been made available by Medicaid or managed care organizations for guidance on reimbursement processes. This highlights an opportunity for Medicaid education to ensure client access to doula services and maintenance of the doula workforce. Doula care is associated with better perinatal outcomes, but structural barriers hinder the ability for low-income families to receive doula care in Florida, creating further health inequities.

---

A doula is a trained non-medical professional that provides physical, emotional, and educational support throughout the perinatal journey. Doula assistance during pregnancy and delivery has been associated with fewer cesarean deliveries, shorter labors, less birth complications, and higher breastfeeding initiation rates (Bohren, Hofmeyr, Sakala, Fukuzawa, & Cuthbert, 2017; K. J. Gruber, S. H. Cupito, & C. F. Dobson, 2013; Hans, Edwards, & Zhang, 2018). Doula clients report higher rates of satisfaction with their birth experiences and increased beliefs in their own abilities to influence birth outcomes through behavior change (Kenneth J Gruber, Susan H Cupito, & Christina F Dobson, 2013). Additionally, doula services have been linked to improved birth outcomes among women experiencing disparate rates of maternal mortality (MM) and severe maternal morbidity (SMM), specifically those in lower socioeconomic brackets or who are Medicaid-eligible (Alexandria et al., 2023; Katy Backes Kozhimannil, Hardeman, Attanasio, Blauer-Peterson, & O’brien, 2013; Ramey-Collier et al., 2023). Importantly, the inclusion of community-based, culturally competent doula support has been associated with fewer maternal complications among socially disadvantaged women facing disproportionately high rates of MM/SMM (Everson, Cheyney, & Bovbjerg, 2018; Gordon et al., 1999; K. J. Gruber et al., 2013; Hans et al., 2018; Mosley et al., 2021).

Disparities in adverse maternal health outcomes among pregnant women at highest risk (e.g., those who are Medicaid-eligible and/or who are non-Hispanic Black) are further complicated by disparate access to beneficial services. When compared to White and privately insured women, Black and publicly insured women are almost twice as likely to *want* but not *have* doula care (Katy B Kozhimannil et al., 2014). Currently, out-of-pocket costs for comprehensive doula services can range from $800 to greater than $2,500 across the United States (Mehra, Cunningham, Lewis, Thomas, & Ickovics, 2019; Safon, McCloskey, Ezekwesili, Feyman, & Gordon, 2021; Weiss, 2022). Further evidencing cost as a barrier to services, most doulas surveyed in previous research have reported only accepting cash payment for their services (Lantz, Low, Varkey, & Watson, 2005). This “self-pay” model may be contributing to the racial disparities noted among those who utilize doula services. Nationally, Medicaid financed 41% of births in 2021(KFF, 2022a). Medicaid financed 45.5% of births in the state of Florida in 2021; with 64.9% accounting for Black birthing people (Florida Health Charts, 2021).

The White House released a “Blueprint for Addressing the Maternal Health Crisis” in June of 2022, proposing expanding access to doula care for Medicaid enrollees and including postpartum coverage as strategies to address racial disparities in maternal mortality and morbidity. Efforts to expand access to doula care for Medicaid enrollees have since steadily increased nationwide (Chen, 2022). Fourteen states and Washington D.C. are actively providing Medicaid coverage for doula services, 11 states are in implementation phases (e.g., legislation has passed, doula services have been incorporated into state budgets), and 13 states are taking action related to doula care (e.g., state certification for doulas, pilot programs) (Chen, 2022).

Of all states actively providing Medicaid coverage for doula services, Florida is the only one that does not utilize a State Plan Amendment (SPA). Instead, Florida includes doula care as an optional expanded benefit that Managed Care plans can include (Gebel & Hodin, 2020; “Health and Human Services”, 2021; Katy B Kozhimannil, Vogelsang, & Hardeman, 2015; Minnesota Department of Human Services, 2020; “Oregon Medicaid reimbursement for doula services”, 2018). Florida’s operationalization of Medicaid reimbursement for doula services warrants analysis, as SPAs ensure greater compliance with federal rules, whereas the use of an expanded benefit means that 1) only 83% of Florida’s Medicaid enrollees in managed care plans can access the benefit, (KFF, 2022b) and 2) each *plan* is permitted to implement the doula benefit rather than following their state’s directive (Robles-Fradet, 2021). In Florida, Medicaid coverage for postpartum services was recently extended from 60 days to 365 days; however, the total number of doula visits allowed (two) was not expanded.

As more than half of all states are currently providing coverage for doula services, in the process of implementation of these services, or proposing legislation for such coverage, understanding doula perspectives regarding reimbursement is critical to being able to serve more families in need (Bey, Brill, Porchia-Albert, Gradilla, & Strauss, 2019; Chen, 2022), such as those with lower incomes.

## A Reproductive Justice Approach

Our conceptualization for this study relies on a reproductive justice approach, which advocates for women having the right to choose if, when, and how to become a parent (Pollitt, 2014; L. Ross, 2020; L. J. Ross, 2017; B. Sundstrom et al., 2019; Beth Sundstrom et al., 2019). An important tenet of the framework is the consideration of various social and economic factors that may support or prohibit a woman’s ability to carry or deliver a baby in a safe and healthy environment (Gurr, 2014; Kluchin, 2016; Pollitt, 2014; L. Ross, 2014). Such considerations highlight how the systemic oppression of Black, Indigenous, and poor women has continued through the intentional limiting of reproductive options in these communities over time (L. Ross, 2020; L. J. Ross, 2017). A reproductive justice approach allows for the intersections of race, gender, and socioeconomics to be explored with the intent of questioning how social systems have influenced reproductive decisions in these communities (L. J. Ross, 2017). Because of the time doulas spend with clients throughout the perinatal period, they have a unique opportunity to identify and illuminate such intersectional identities, providing access to important social programs.

## The Theoretical Domains Framework

The Theoretical Domains Framework (TDF) provides a useful ‘first step’ toward recognizing and classifying barriers and facilitators that impact implementation of services for Medicaid recipients (Cane, O’Connor, & Michie, 2012). By structuring qualitative data as barriers and facilitators, the TDF permits for targeted intervention (Guerra, Lambe, Manolova, Sadler, & Sheehan, 2022). Comprised of 14 validated domains (knowledge; skills; social/professional role and identity; beliefs about capabilities; optimism; beliefs about consequences; reinforcement; intention; goals; memory, attention, and decision processes; environmental context and resources; social influences; emotion; and behavioral regulation) to help organize social, cognitive, environmental, and affective influences on one’s behavior, the TDF has previously been applied in qualitative health research to determine healthcare professionals’ perceptions across a variety of settings (Guerra et al., 2022; Pearse, Keogh, Rickard, & Fung, 2021). Previous work has documented the TDF’s usefulness for the synthesis of qualitative evidence related to ‘policy-urgent’ questions (Shaw, Holland, Pattison, & Cooke, 2016), making Florida’s Medicaid reimbursement process an area ripe for exploration.

## Materials and Methods

### Interviews and Focus Groups

Utilizing the framework for conducting TDF research by Atkins et al. (Atkins et al., 2017) interviews and focus groups were chosen for the study design to illuminate Florida doulas’ subjective experiences with the Medicaid reimbursement process. We received exempt status from two Institutional Review Boards. Convenience and snowball sampling were used for recruitment. Flyers were shared with community partners and distributed within their networks. Eligibility required that a participant serve as a birth doula with experience in Florida and have served at least two families in the past year. Interview and focus group guides focused on doulas’ experiences with Medicaid, the reimbursement process, and what they may have heard from other doulas experiences with the process. Interviews were used to triangulate focus group findings, and to provide more convenient scheduling options for participants needing to reschedule due to client births. Informed consent was collected for all participants. Interviews and focus groups were conducted in spring and summer of 2022. The first author, a qualitative health communication scholar, facilitated most sessions. The third author, after proper training, facilitated some sessions. Note takers were present in all sessions for memo purposes. Sessions lasted approximately 90 minutes, were digitally recorded, and initially transcribed using AI technology. Members of the research team then cleaned all transcripts by hand to ensure accuracy.

### Data Analysis

#### Phase One – Inductive Coding

For phase one, our team relied upon a general inductive approach to coding, where codes were created based on the data generated (Thomas, 2006). Saturation of data was reached, meaning that as more data were collected, little additional information was revealed, making further data collection counter-productive (Morse, 1995). We utilized the intercoder agreement approach for qualitative interviewing, which aides in improving a coding scheme’s discriminant capability via the unitization of data (Morse, 1995). Transcripts were independently reviewed by all study team members to engage in the open coding phase, where a close, line-by-line reading of all data occurred to look for the emergence of meaningful words and units (Bogdan & Taylor, 1975).

The team then convened to engage in axial coding, where initial codes were generated into broader themes, allowing for codebook creation (Parker, Ang, & Koslow, 2018). The two interview/focus group moderators tested the codebook on a 10% sample of transcripts, which resulted in an intercoder agreement of 92.2% (considered a “very good” level in qualitative research) (Miles & Huberman, 1984). All data were then coded by the first author using NVivo, a qualitative data analysis software.

#### Phase Two – Deductive Coding for the TDF

Following the initial phase of data analysis, we employed the TDF as a secondary lens for viewing all analyzed data. Each primary theme (and their respective codes) were deductively coded into theoretical domains for consideration into appropriate constructs as a directed content analytic technique (Atkins et al., 2017; Hsieh & Shannon, 2005). Team members met twice to discuss final domains and constructs of the TDF.

#### Weekly Stakeholder Meetings and Member Checking

Our research team meets on a weekly basis with community stakeholders (doulas, clinicians, students, public health professionals, key personnel from *The Doula Network/TDN,* etc.) to receive input on current projects and embody the principles of community-based participatory research. Currently, an average of six stakeholders join our virtual calls. We engaged in qualitative member-checking practices of the interview and focus group transcription data, where two stakeholder-participants reviewed data to enhance participant validation. Team meetings, and review of primary themes, served to showcase the complexity of Florida’s Medicaid reimbursement – even for seasoned doulas. Stakeholder participants then engaged other leaders in the field (birth justice advocates, experts in Medicaid reimbursement with multi-state knowledge) to join our meetings to further discuss intricacies surrounding the reimbursement process. These conversations became important for informing the analysis for this manuscript. In sum, through team consultation and engagement with MCO resources and Medicaid literature, approximately nine hours of group meetings were necessary to fill the gaps of Florida’s Medicaid reimbursement process.

## Findings

### Participants

In total, the study sample included 31 doulas serving in Florida. Seven in-depth interviews and seven focus groups were conducted online. The majority (93%) of participants reported serving families who may qualify for government assistance (e.g., Medicaid, WIC) due to economically disadvantaged circumstances. Most who participated (55%) were between 25 and 44 years old, nearly three-quarters (76%) self-identified their race as White, less than one-quarter (17%) self-identified their race as Black, and more than a quarter (28%) self-identified as Hispanic/Latino. For a full demographic breakdown of participants, see Table 1.

**Table 1.** Participant Characteristics.

|  |  |  |
| --- | --- | --- |
| (n=29*) |  |  |
| Characteristic | *Percentage* | *n* |
| **Sex** |  |  |
| Female | 96.6 | 28 |
| Other | 3.4 | 1 |
| **Race** |  |  |
| White | 75.9 | 22 |
| Black | 17.2 | 5 |
| Other | 6.9 | 2 |
| **Ethnicity** |  |  |
| Hispanic/ Latino | 27.6 | 8 |
| Non-Hispanic/ Latino | 72.4 | 21 |
| **Age** |  |  |
| 25-34 | 13.8 | 4 |
| 35-44 | 41.4 | 12 |
| 45-54 | 27.6 | 8 |
| 55-64 | 17.2 | 5 |
| **Education Level (Highest Degree Attained)** |  |  |
| Some college credit/ no degree | 20.7 | 6 |
| Trade/ technical/ vocational training | 10.3 | 3 |
| Associate's degree | 13.8 | 4 |
| Bachelor's degree | 31.0 | 9 |
| Master's degree | 17.2 | 5 |
| Professional degree | 6.9 | 2 |
| **Employment** |  |  |
| Employed for wages | 51.7 | 15 |
| Self-employed | 44.8 | 13 |
| Homemaker | 3.4 | 1 |
| **Relationship Status** |  |  |
| Married | 51.7 | 15 |
| In a relationship but not married | 3.4 | 1 |
| Never married | 17.2 | 5 |
| Separated | 10.3 | 3 |
| Divorced | 17.2 | 5 |
| **Household Structure** |  |  |
| Female (single) head of household | 41.4 | 12 |
| Dual 2 parent | 48.3 | 14 |
| Dual 2 other- relative/ kinship care | 3.4 | 1 |
| Other | 3.4 | 1 |
| Decline to answer | 3.4 | 1 |
| **Household Income** |  |  |
| \$10,000- \$19,999 | 3.4 | 1 |
| \$20,000- \$29,999 | 13.8 | 4 |
| \$30,000- \$39,999 | 6.9 | 2 |
| \$40,000- \$49,999 | 10.3 | 3 |
| \$50,000 and up | 58.6 | 17 |
| Decline to answer | 6.9 | 2 |

\*31 participants completed interviews or focus groups, but N=29 for demographic information because 2 participants did not complete the demographic questionnaire.

Doulas described their current services and profile, allowing participants to be grouped into one of four categories: (a) currently accepting Medicaid clients (n=5; 16%), (b) not accepting Medicaid clients but would be able to if they so desired (n=10; 32%), (c) not accepting Medicaid clients due to their service being tied to a grant- or community-funded program (n=12; 39%); or (d) about to begin accepting Medicaid clients/in the process of being certified to accept Medicaid clients (n=4; 13%). Notably, review of all coding resulted in participant groups being combined for thematic representation of the Medicaid reimbursement process, as saturation was reached for each category. Doula opinions formed based on other doulas’ accounts they had heard (word of mouth or electronic word-of-mouth) were used to validate the data provided by doulas currently experiencing the Medicaid reimbursement process (Martin, 2017).

## Results

Data analysis highlighted several barriers and minimal facilitators for doulas regarding Florida Medicaid implementation efforts. The barriers, specifically, serve to showcase the complexity of the Medicaid process. Barriers for doulas who are currently engaged with the Medicaid process, and those who are actively avoiding it, were housed within five primary themes: reimbursement rates/doula livelihood; reimbursement process; doula autonomy; certification criteria or Medicaid requirements viewed as unfair/unrealistic; and information gaps/misinformation. Facilitators for doulas currently engaged with the Medicaid process included national structures to help streamline processes and a desire to help communities. Findings are described below with thematic summaries, additional participant quotes, and domains and constructs relevant to the TDF provided in Tables 2 and 3. Domains related to the TDF are included in italic text/parentheses.

**Table 2.**
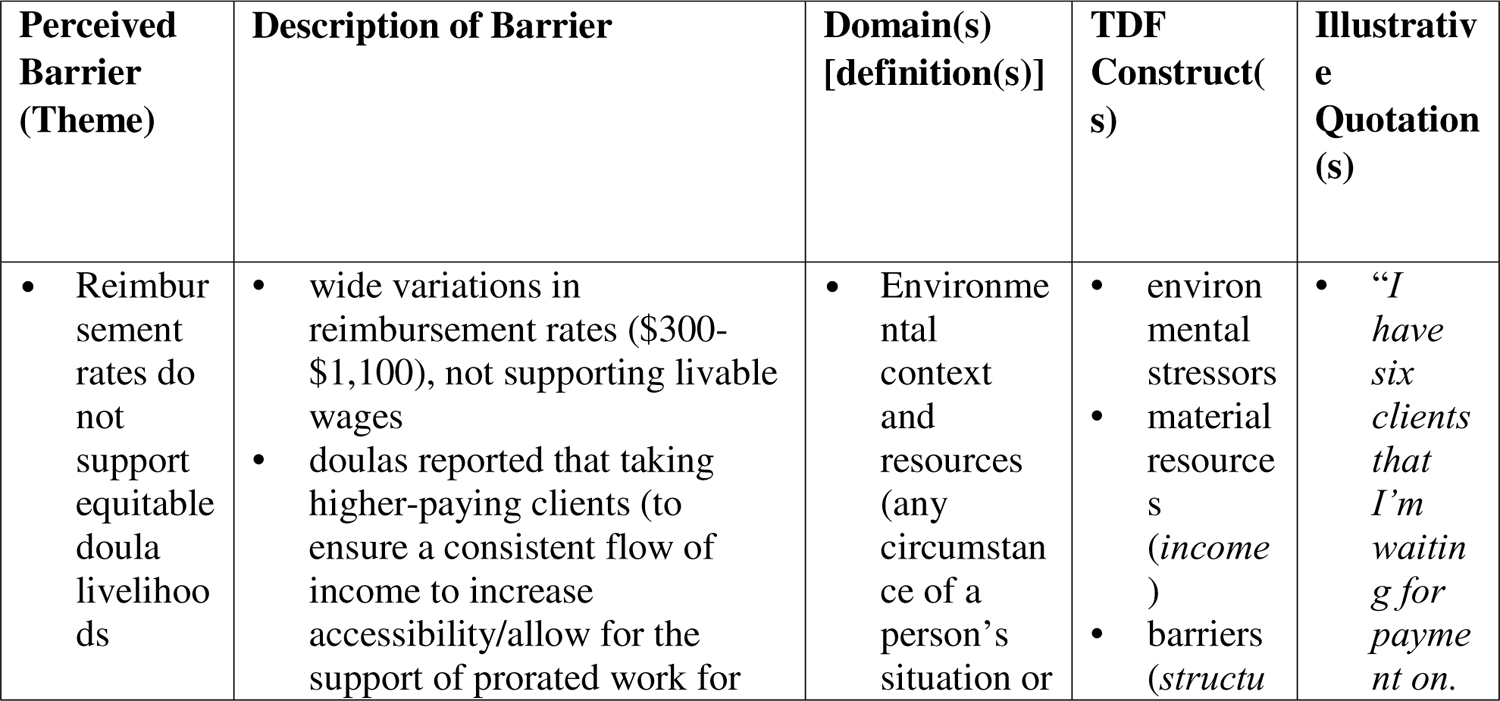
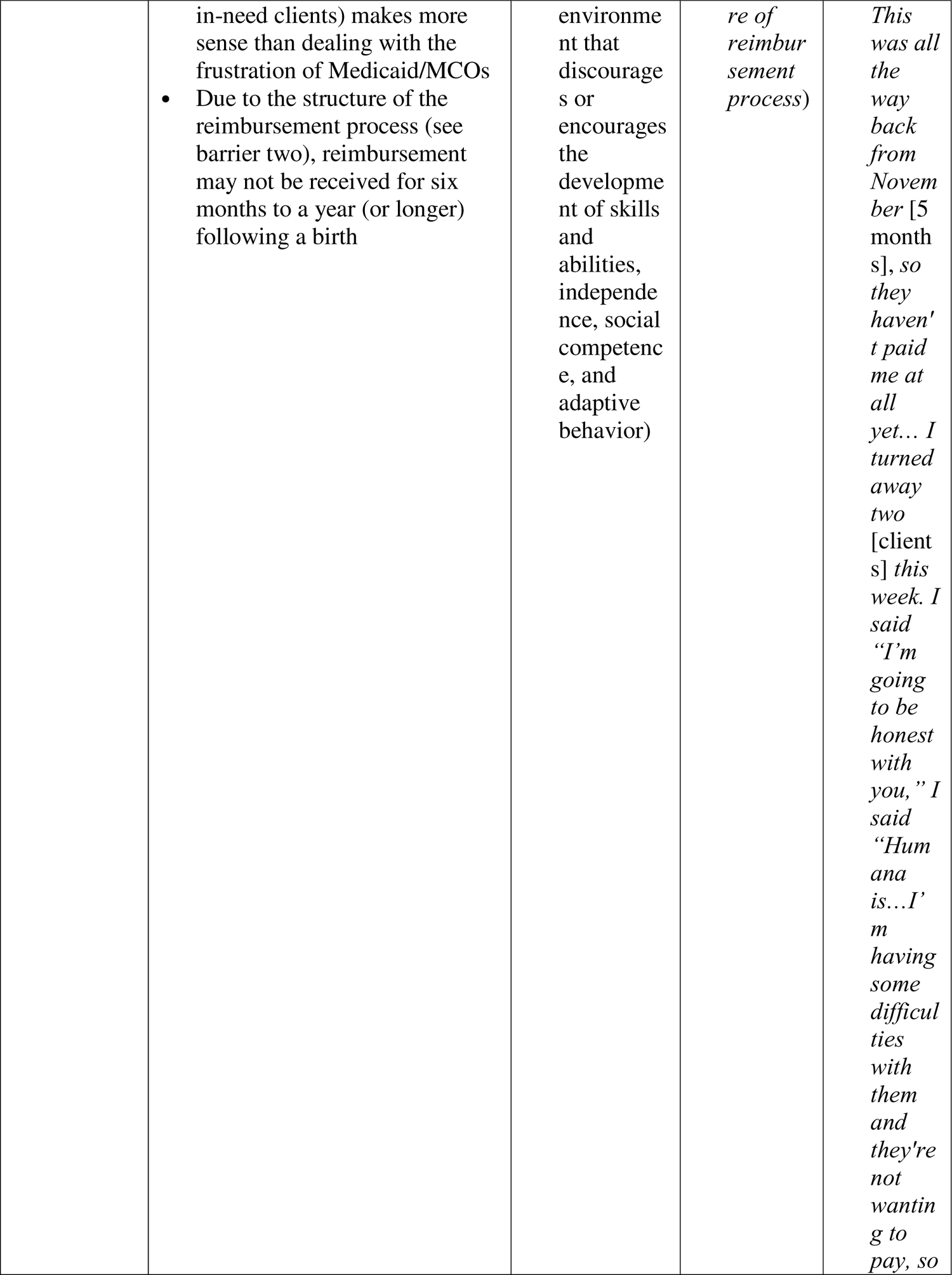
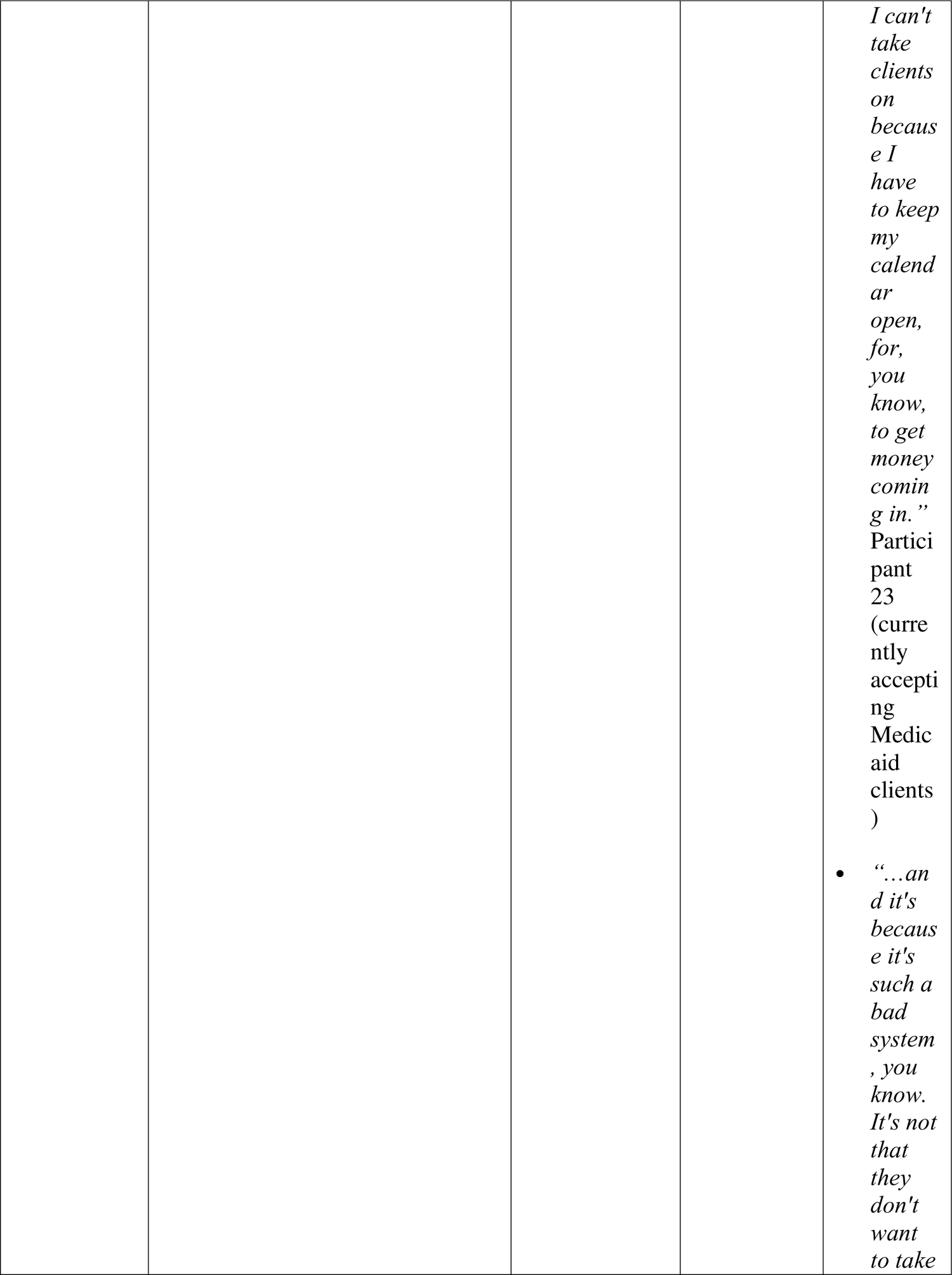
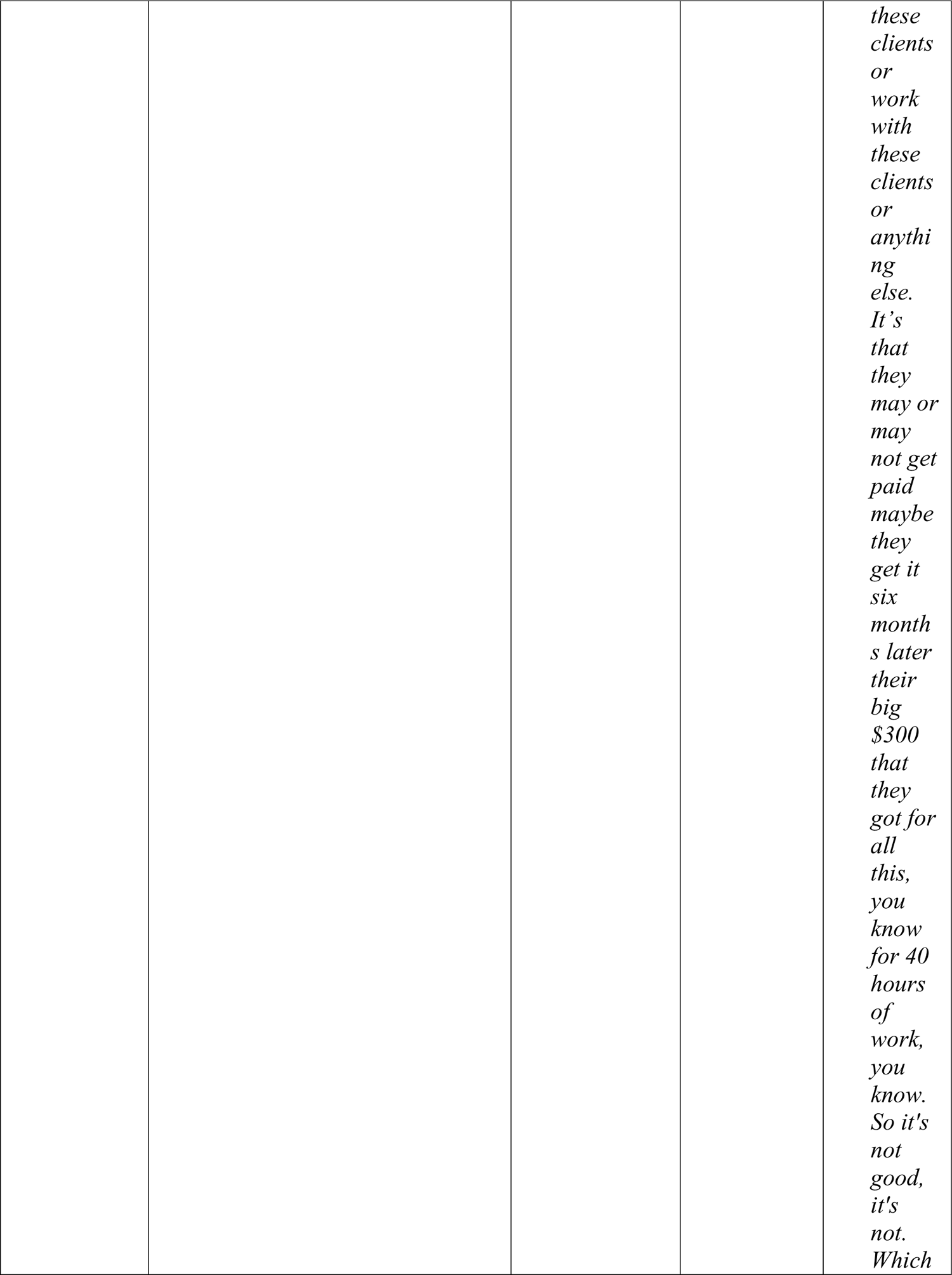
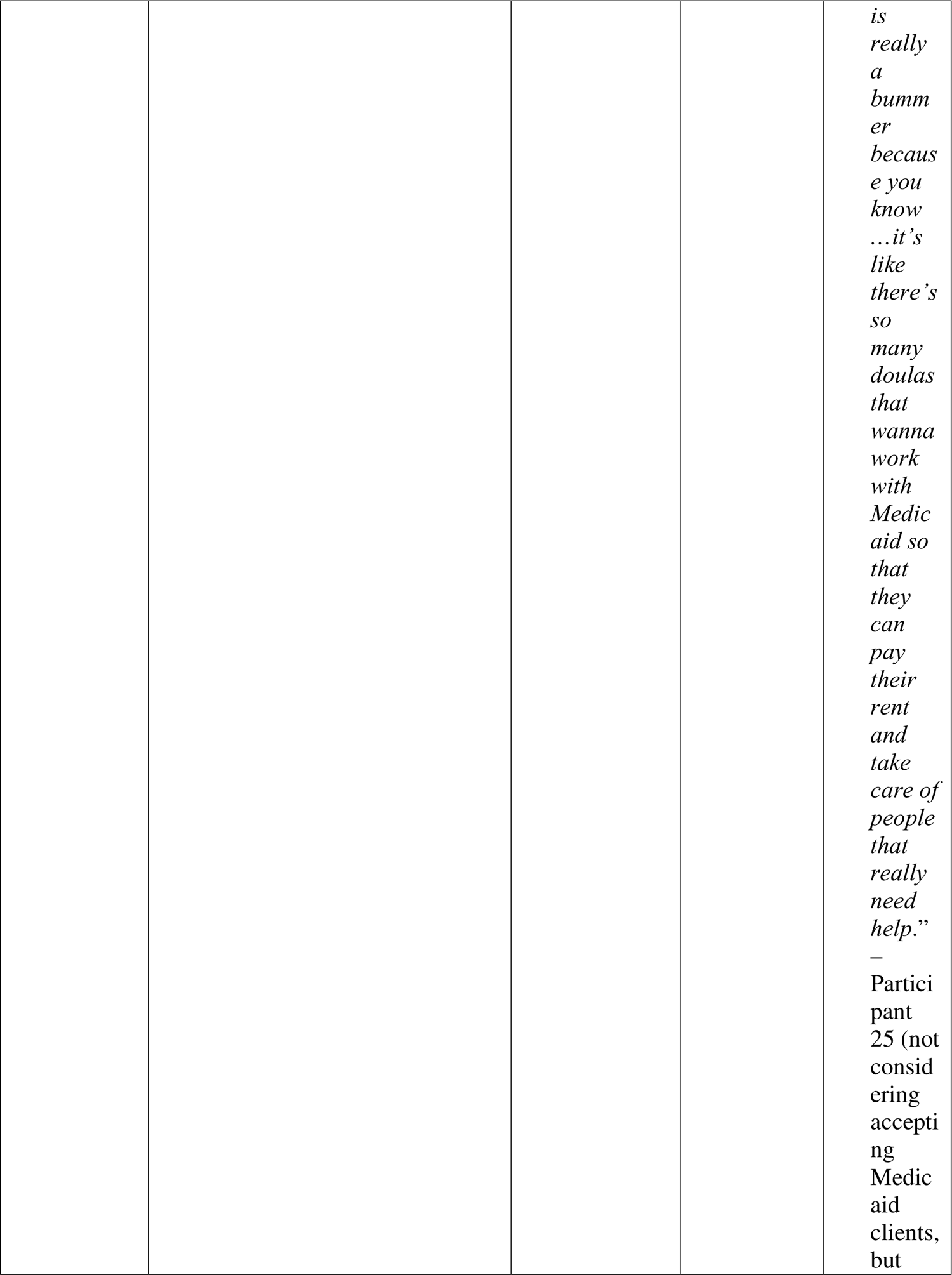
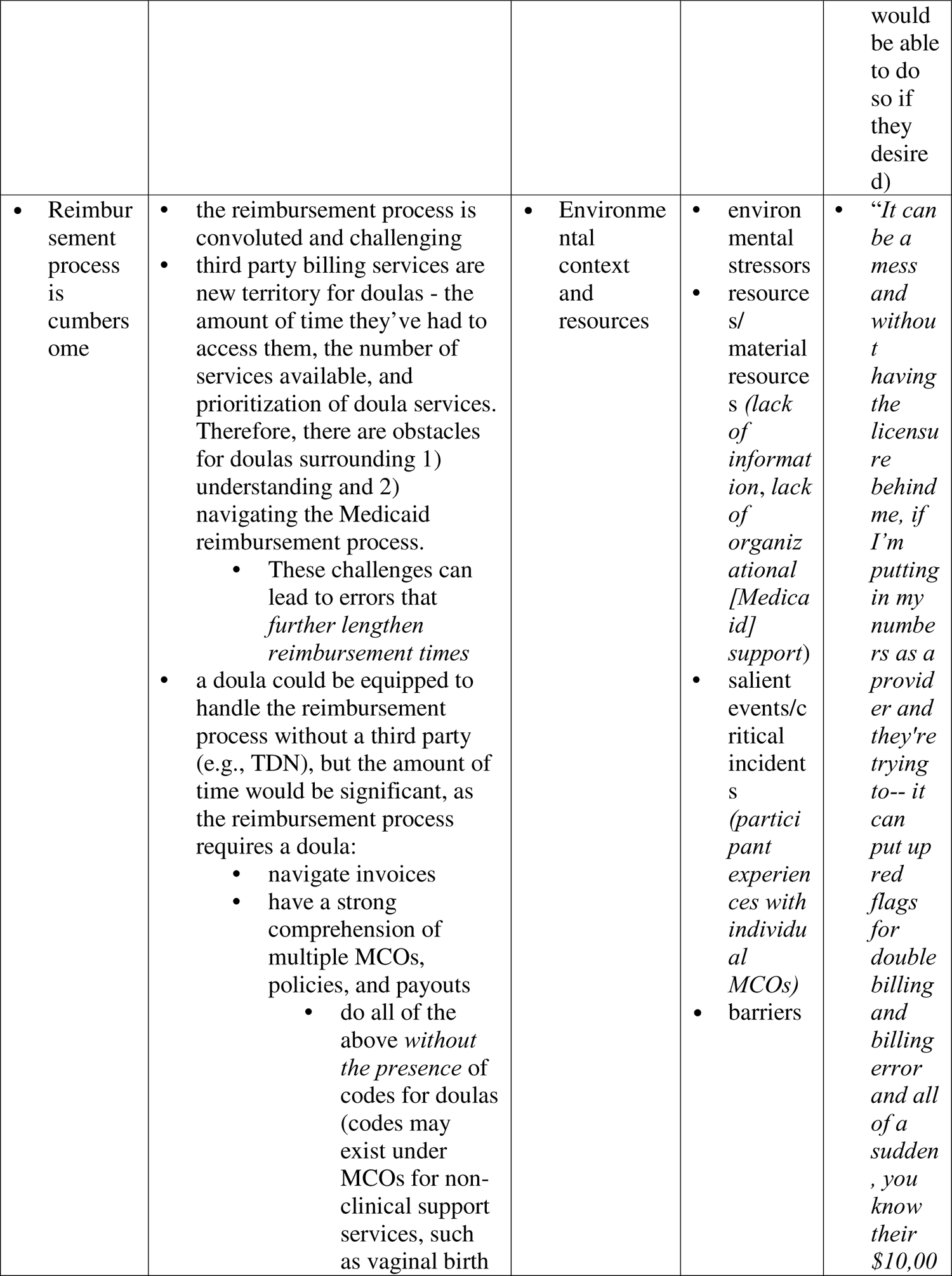
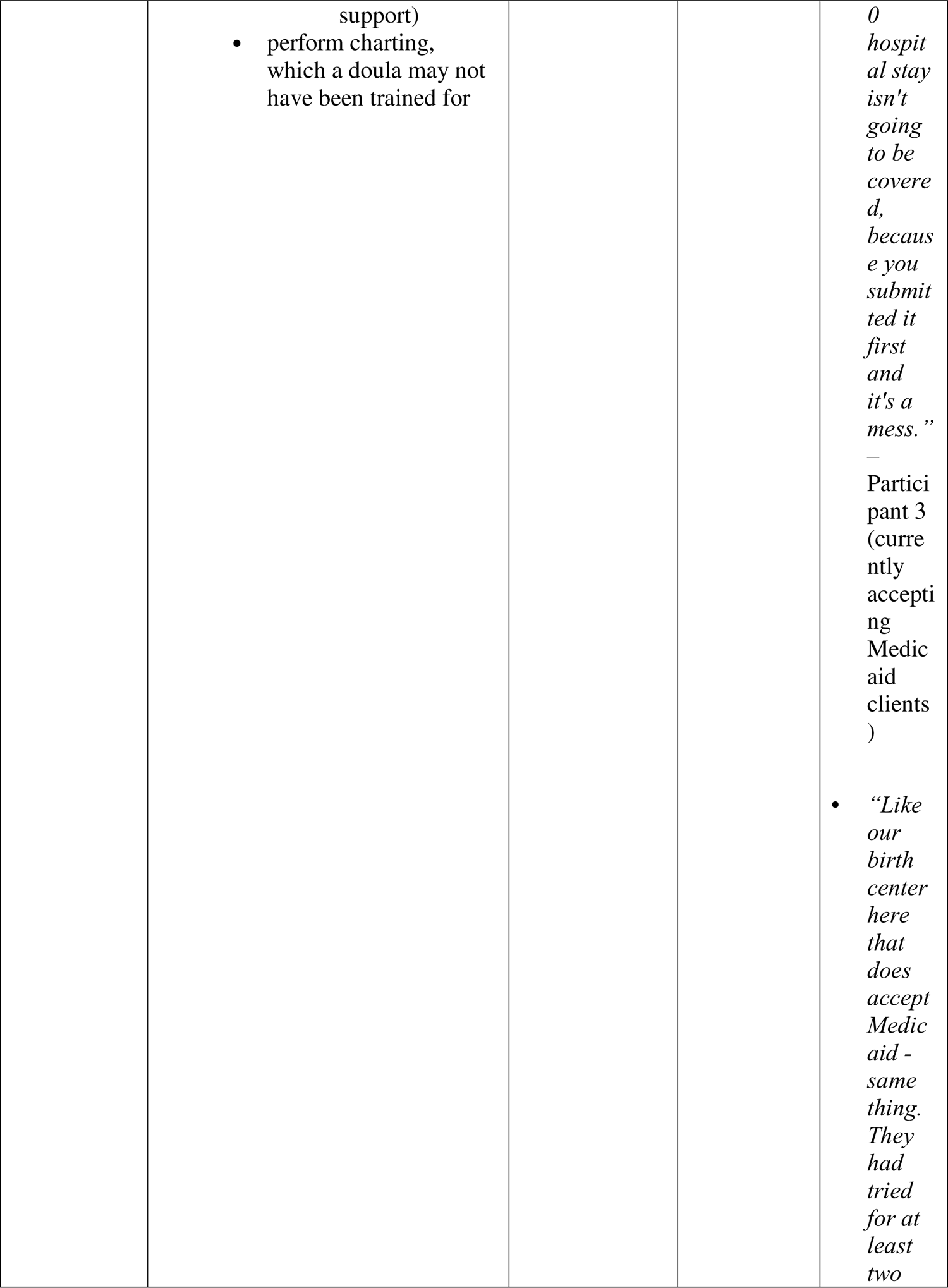
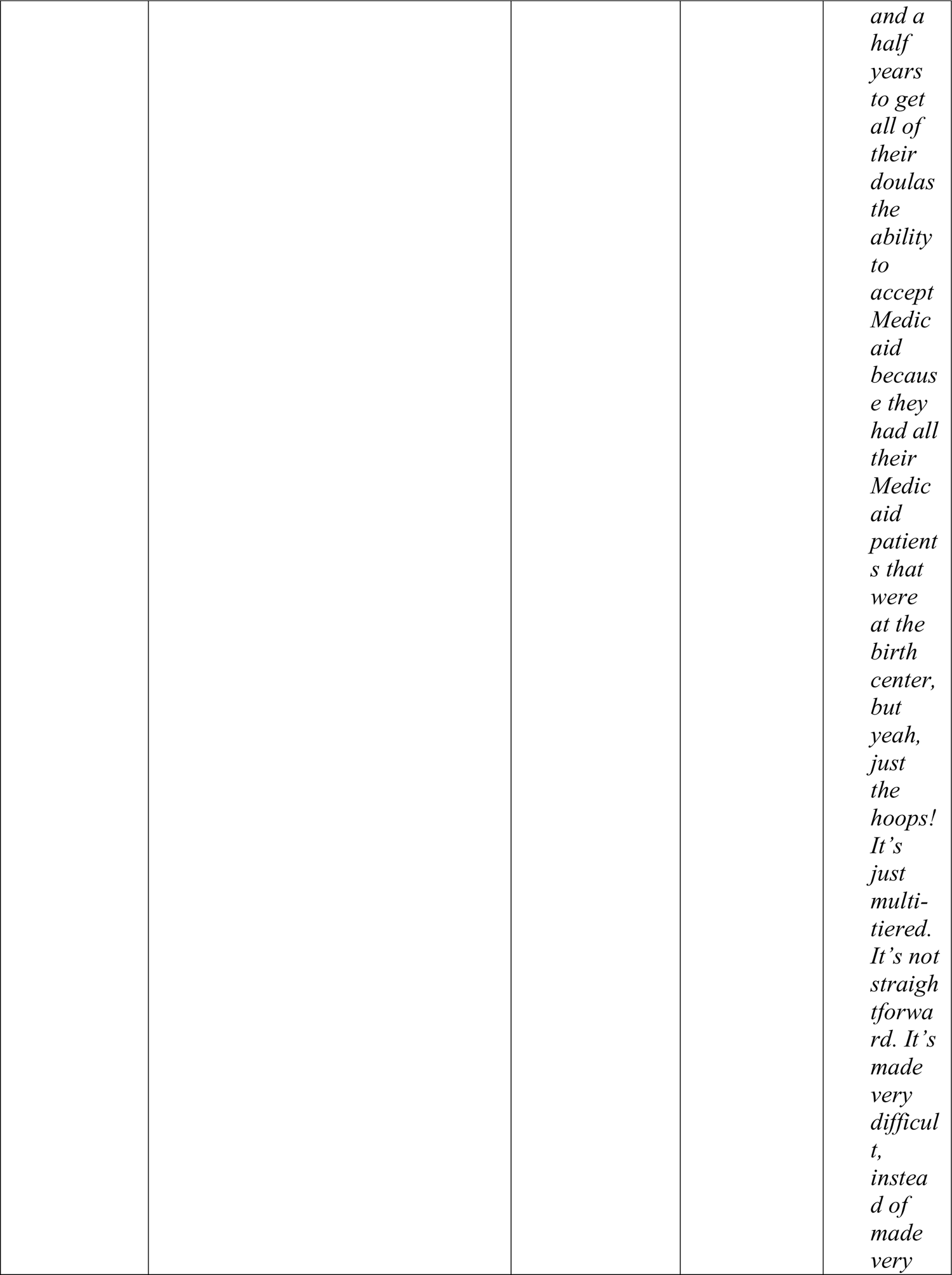
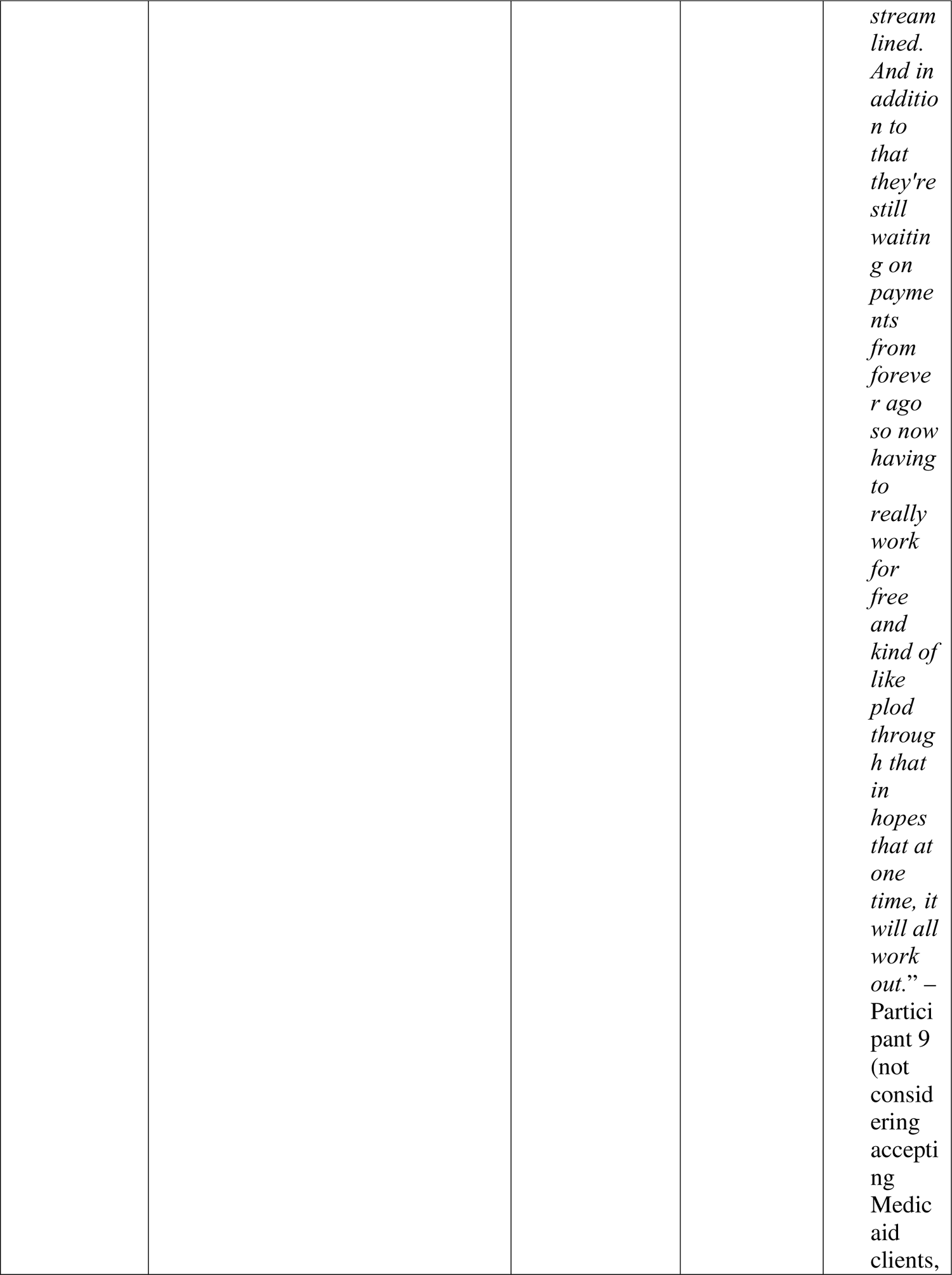
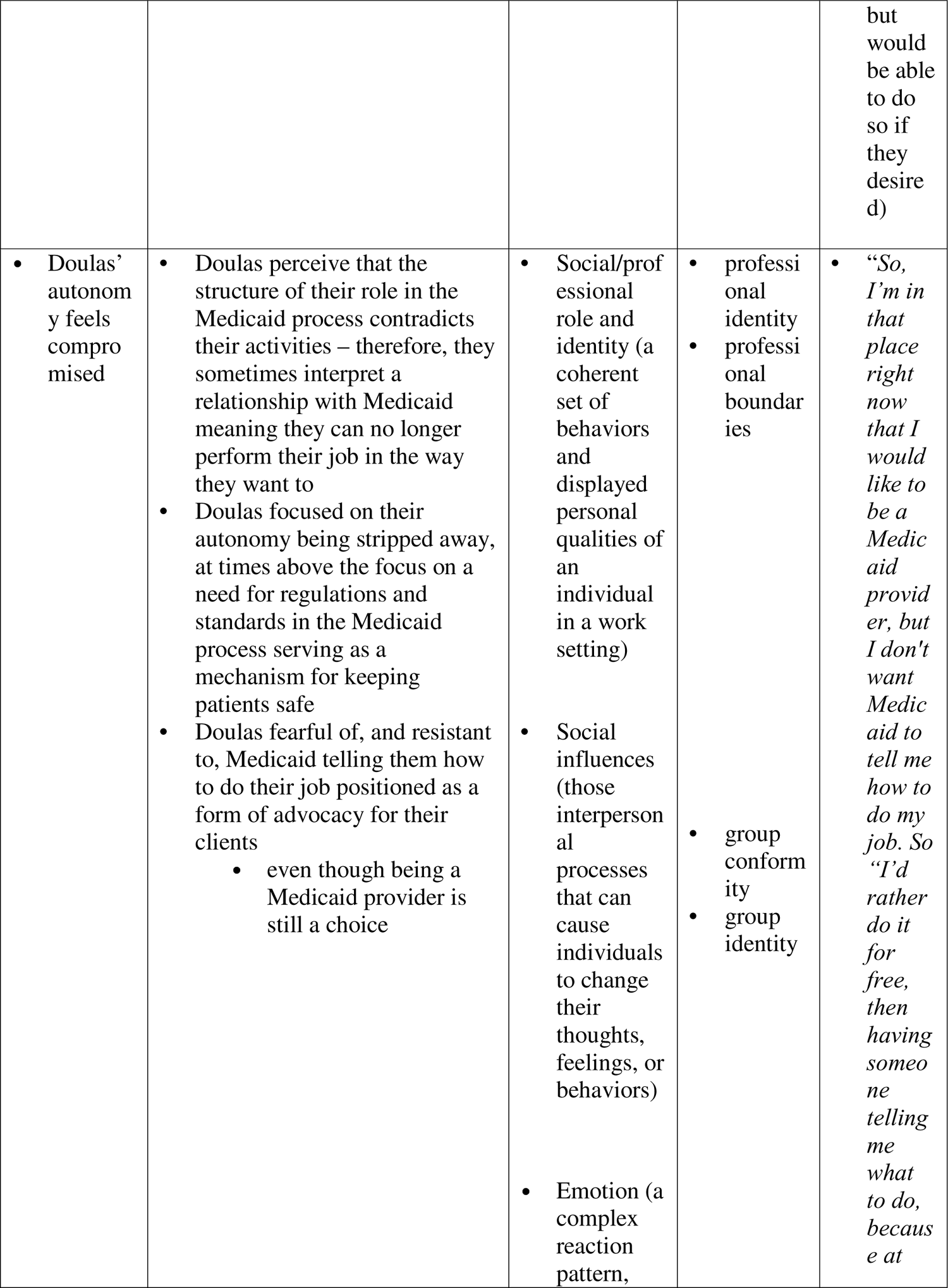
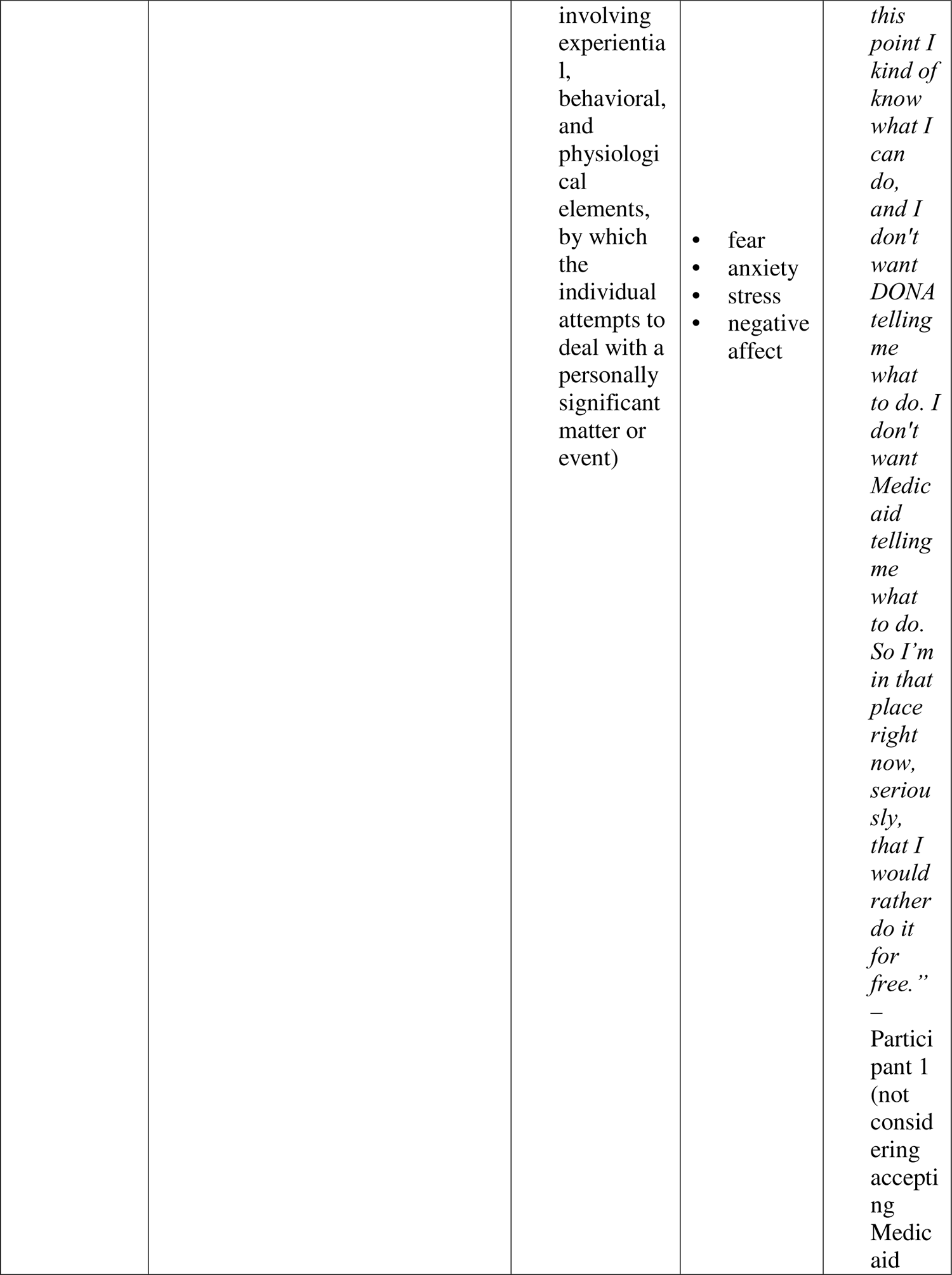
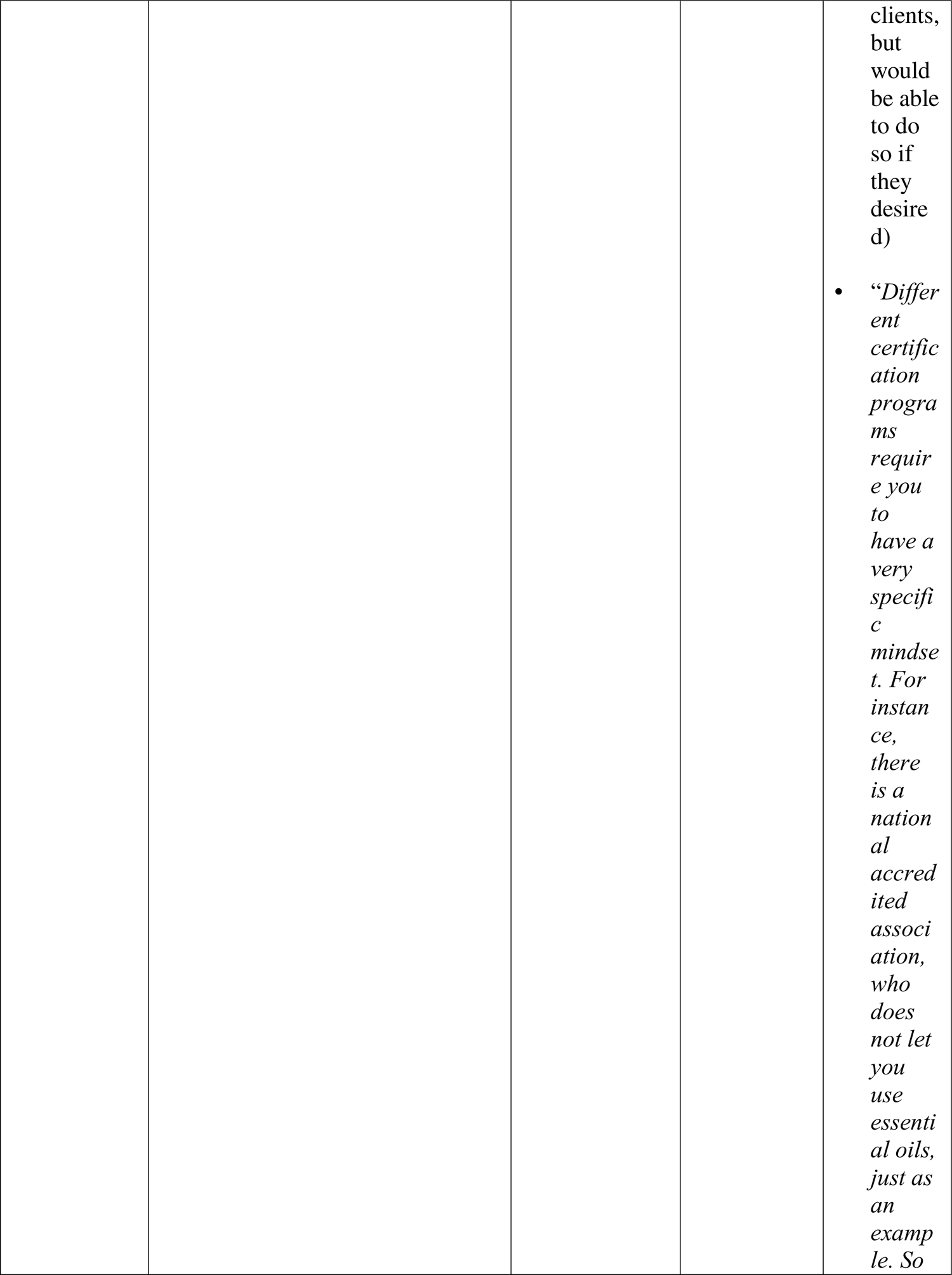
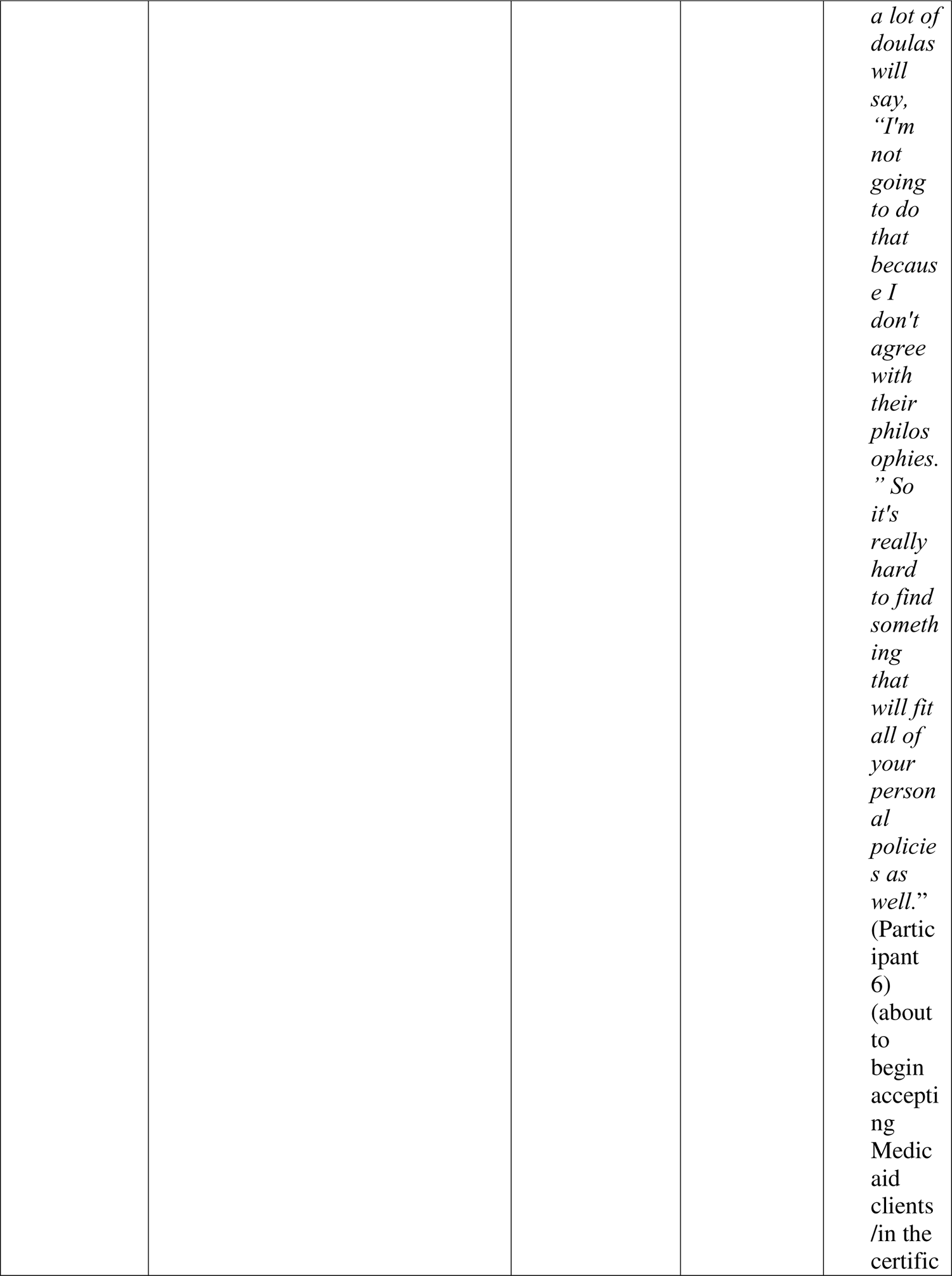
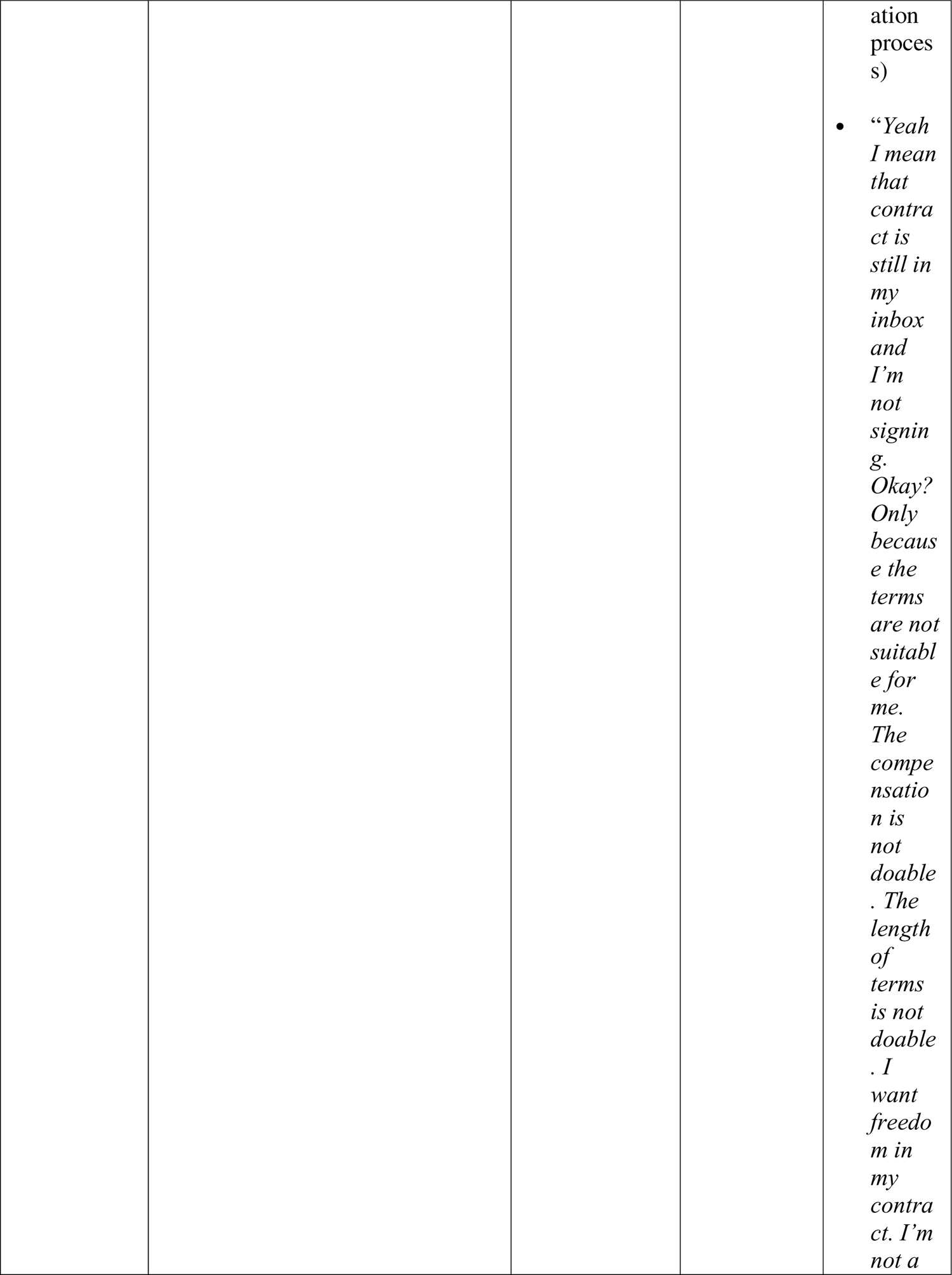
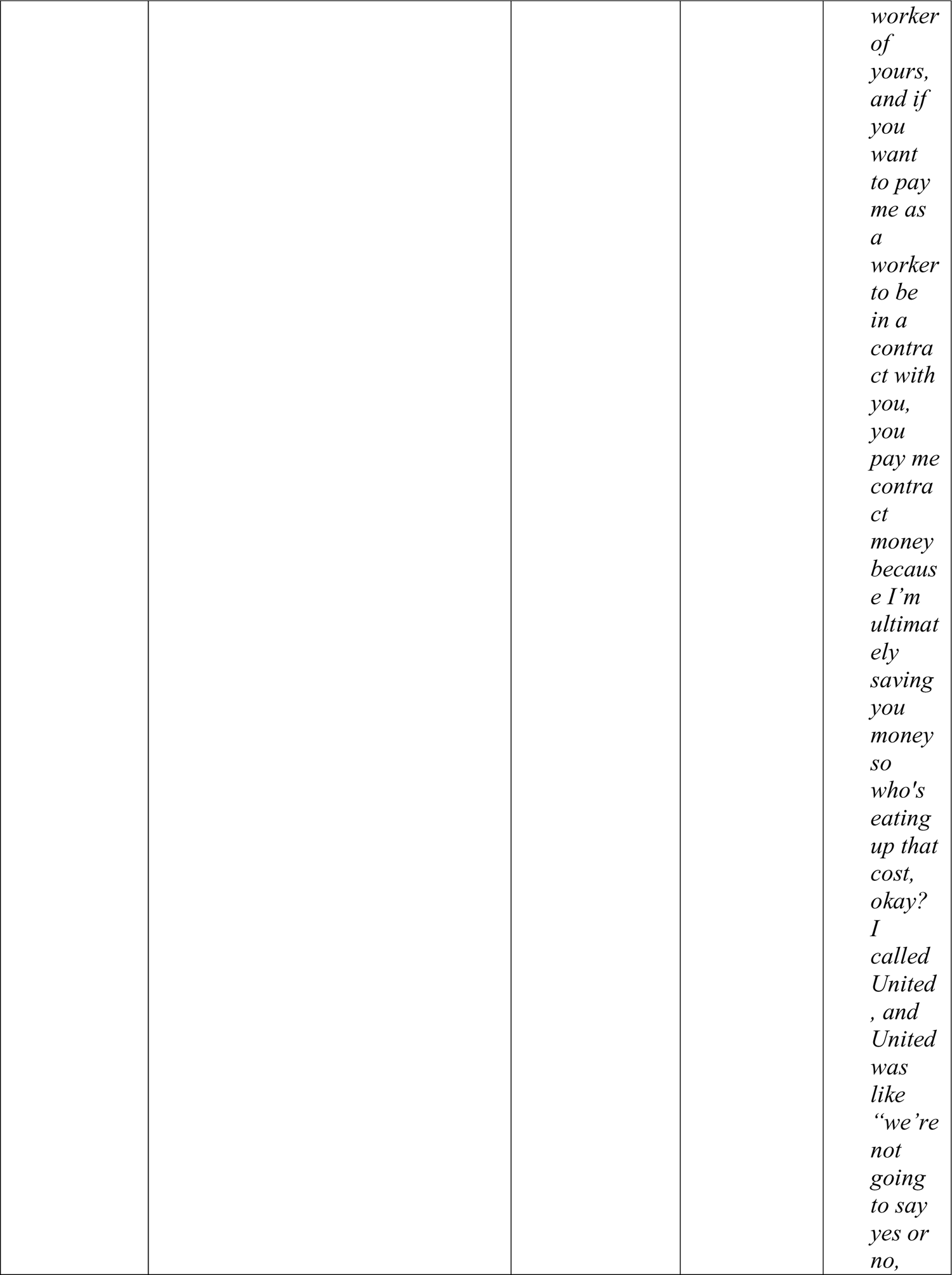
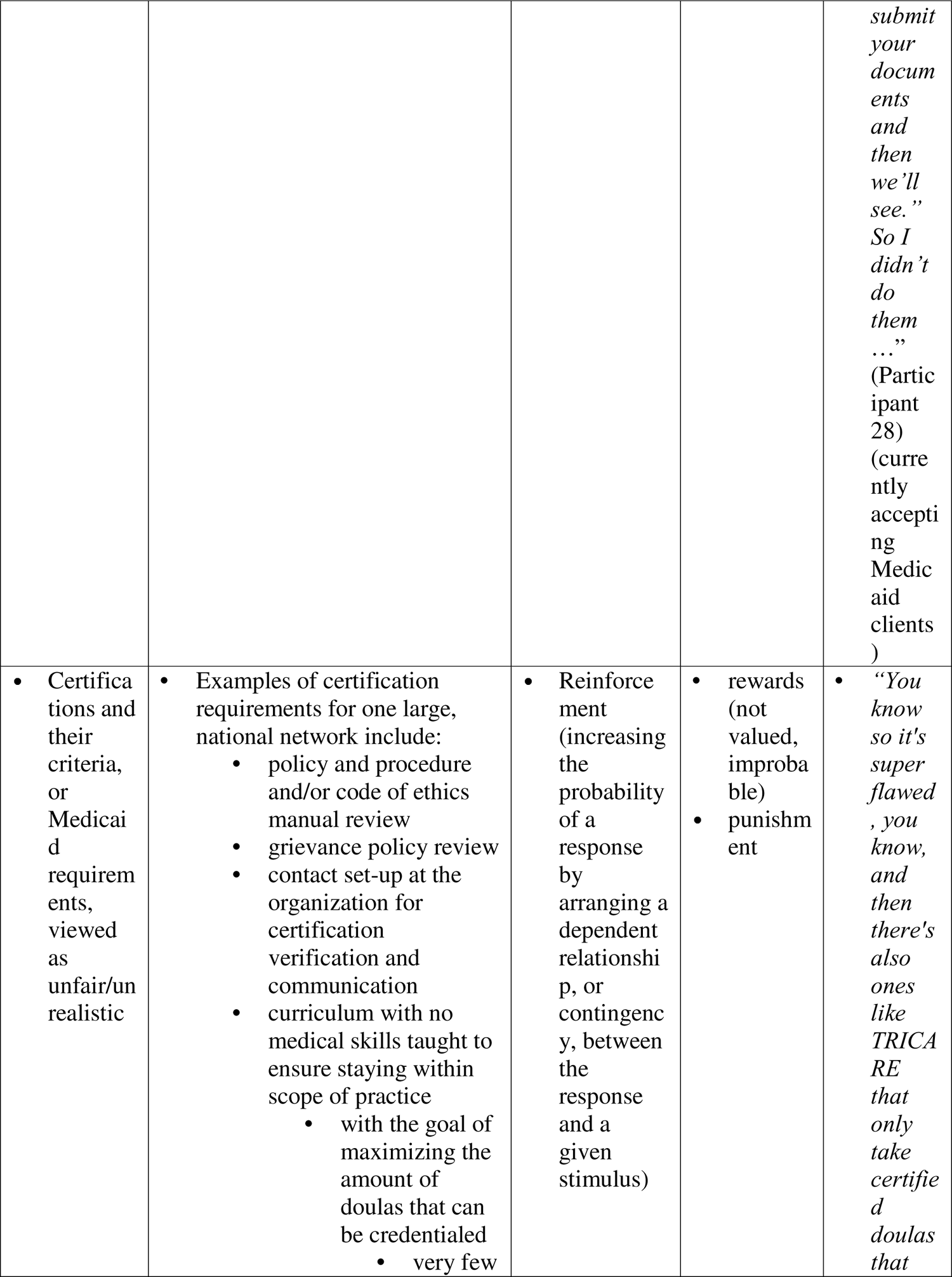
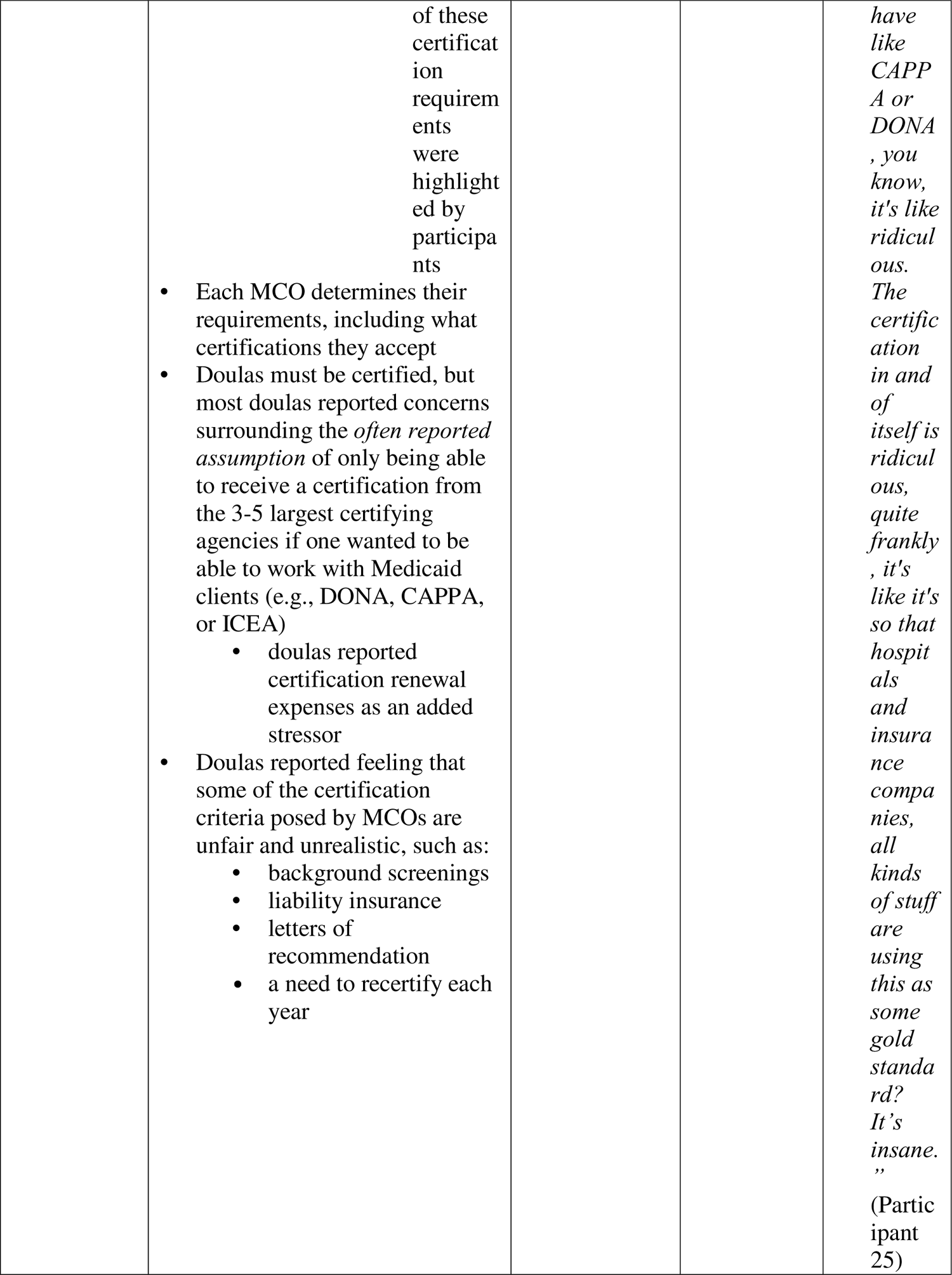
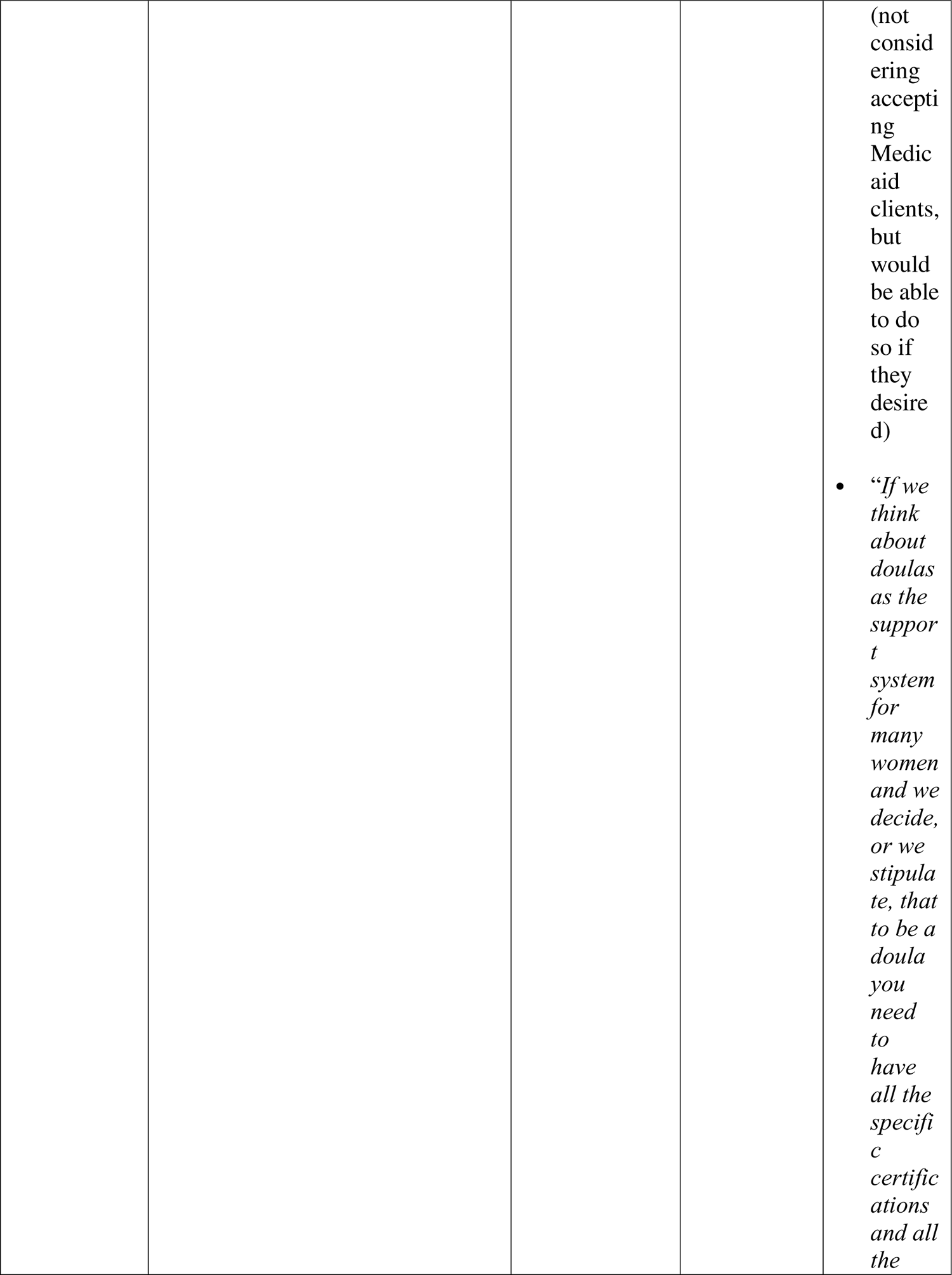
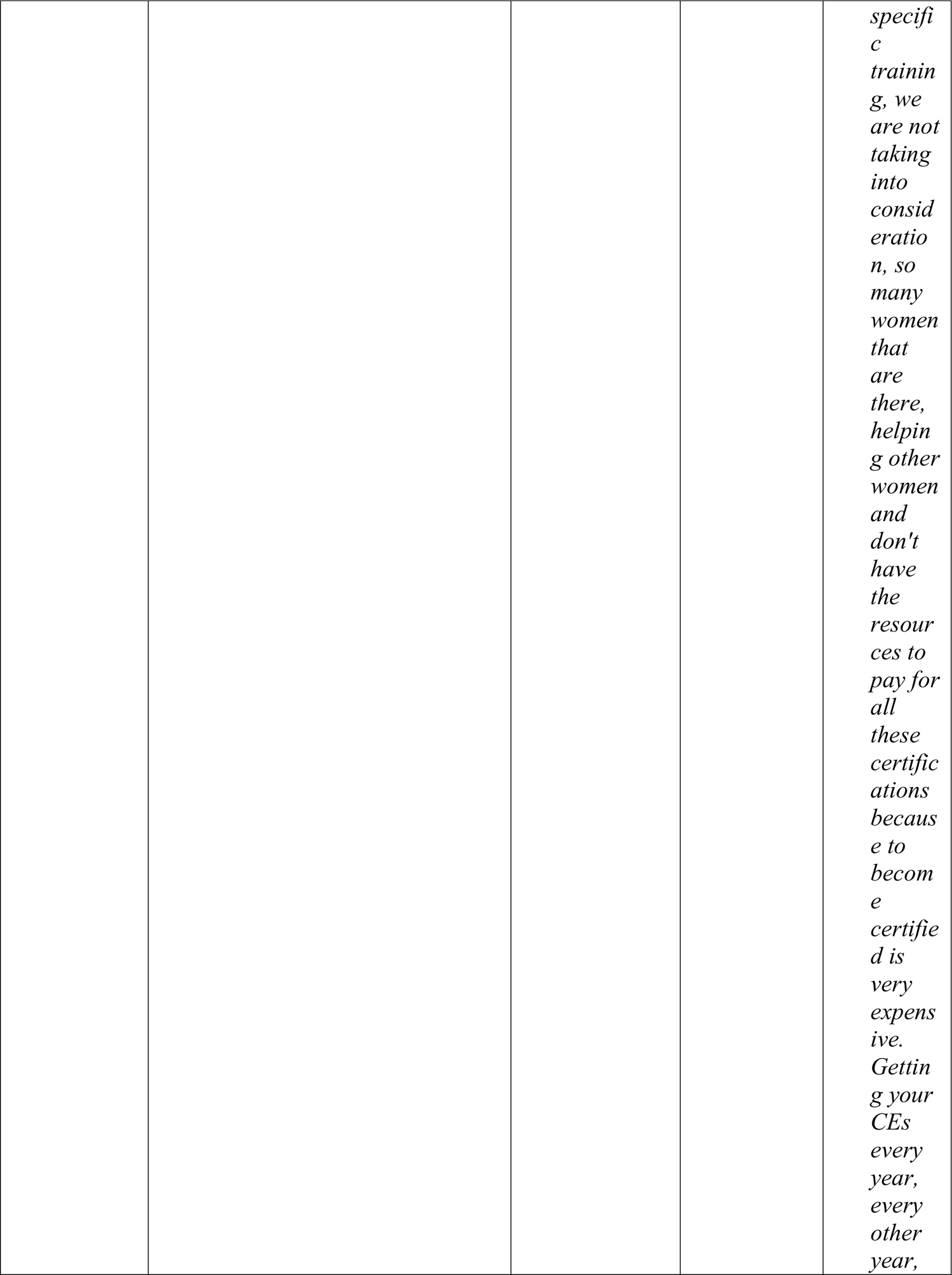
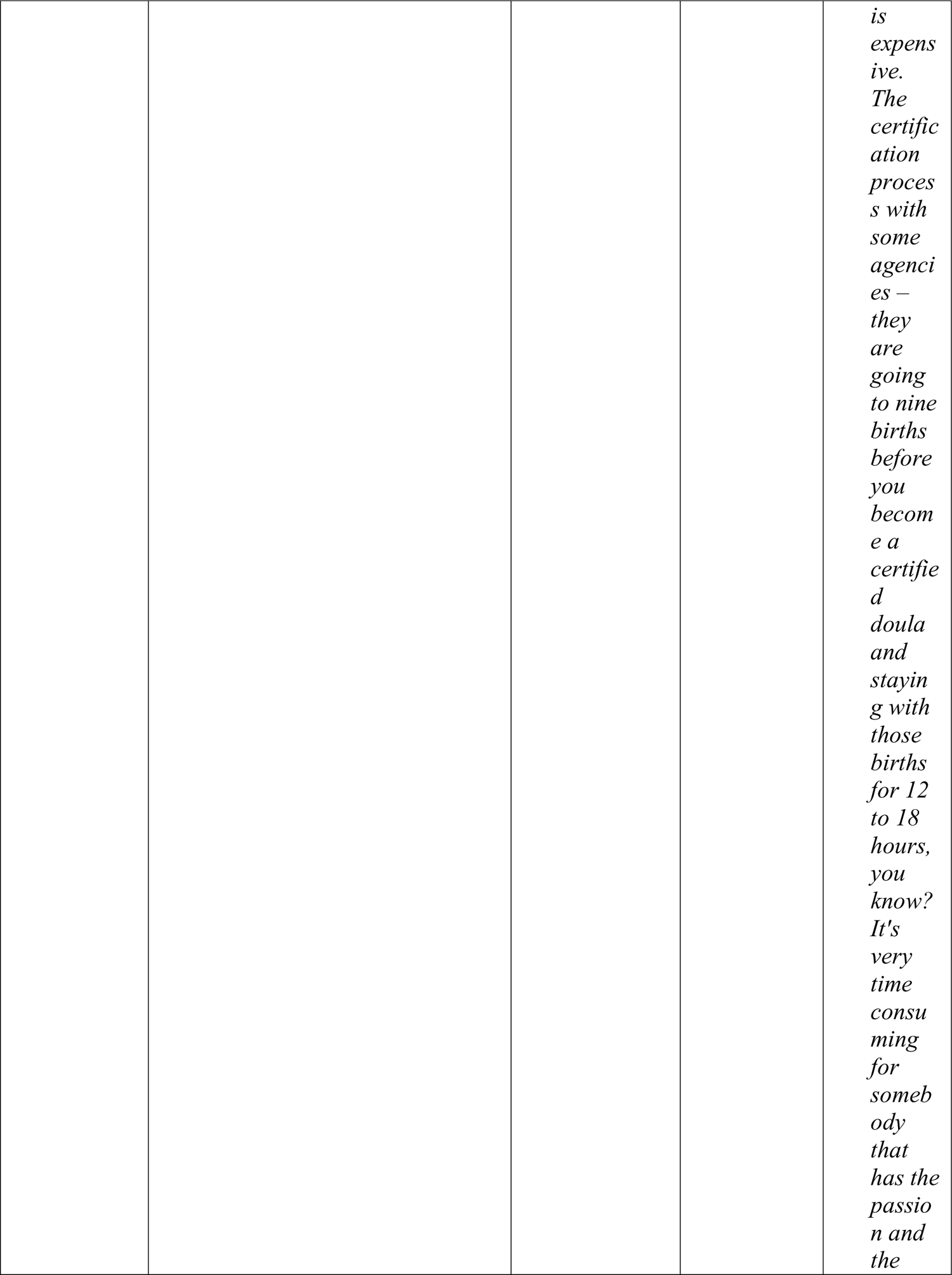
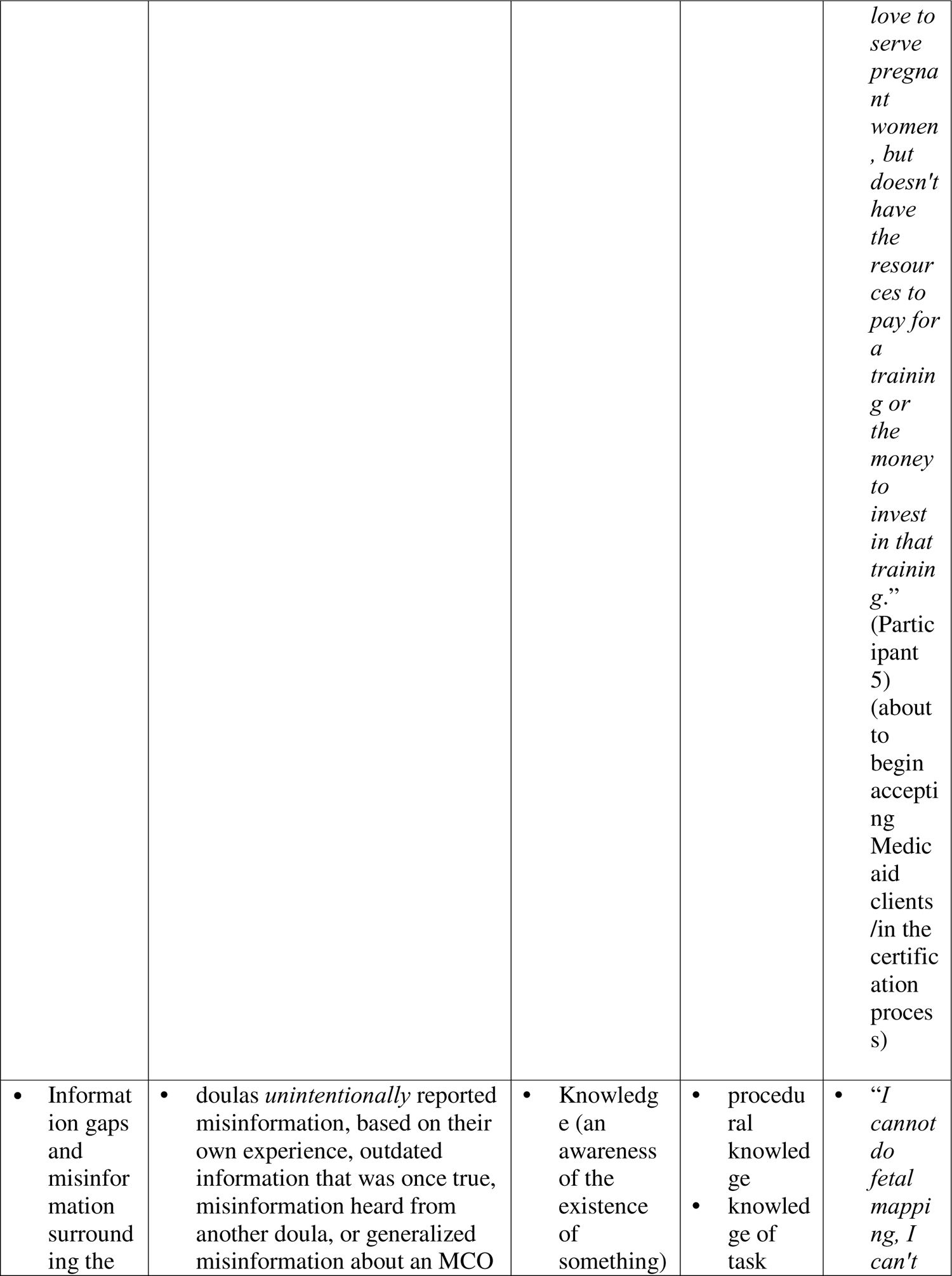
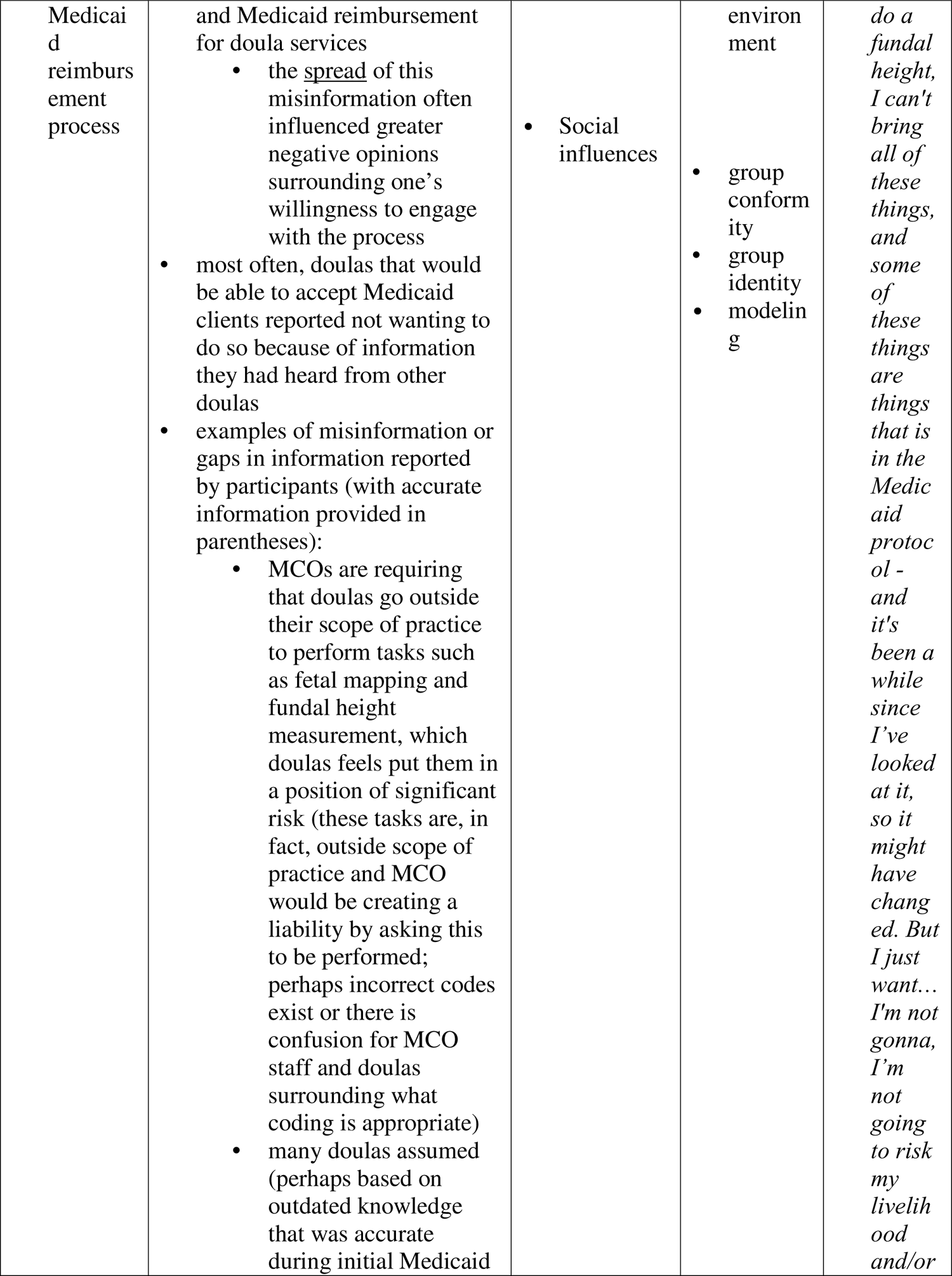
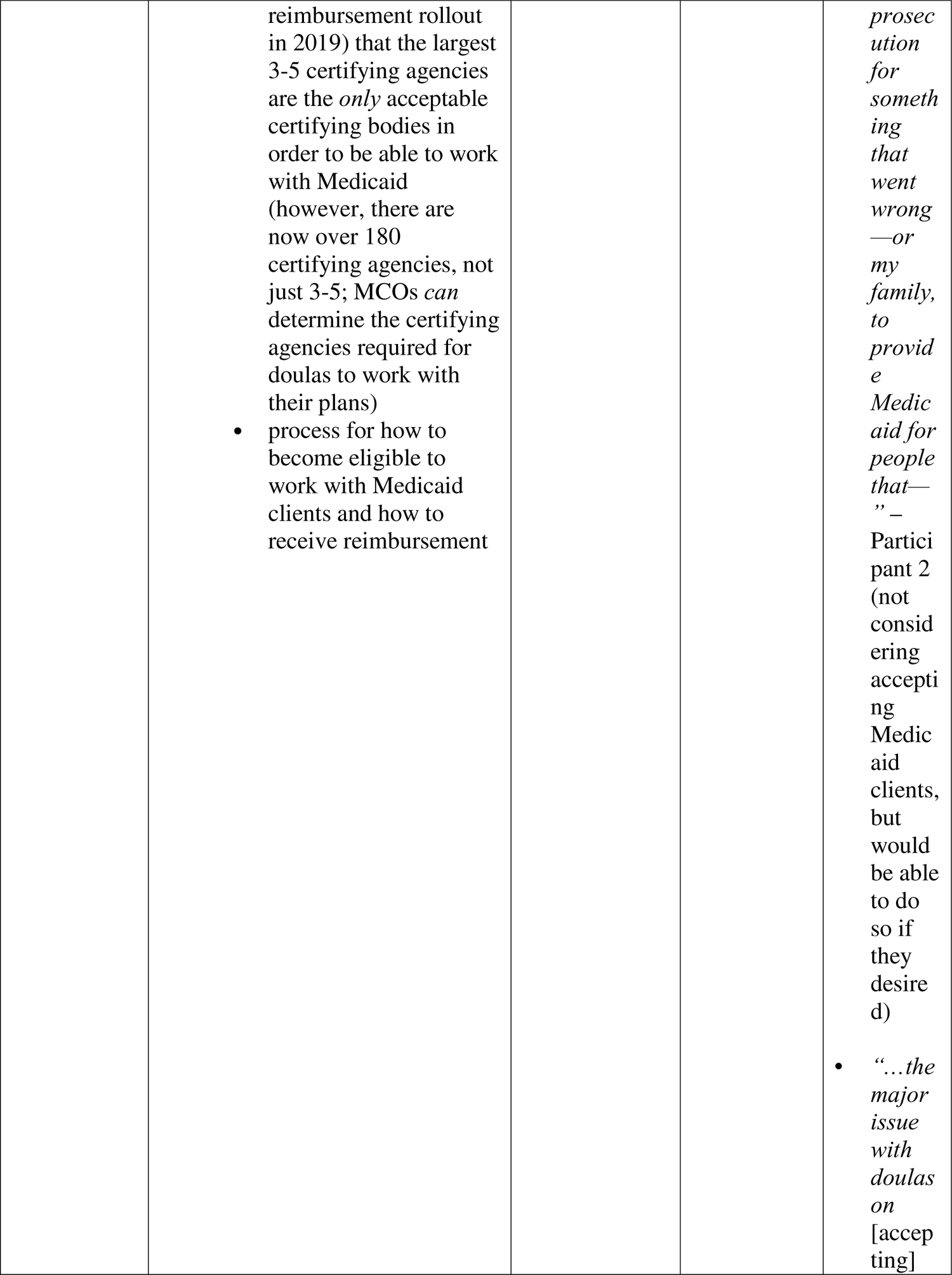
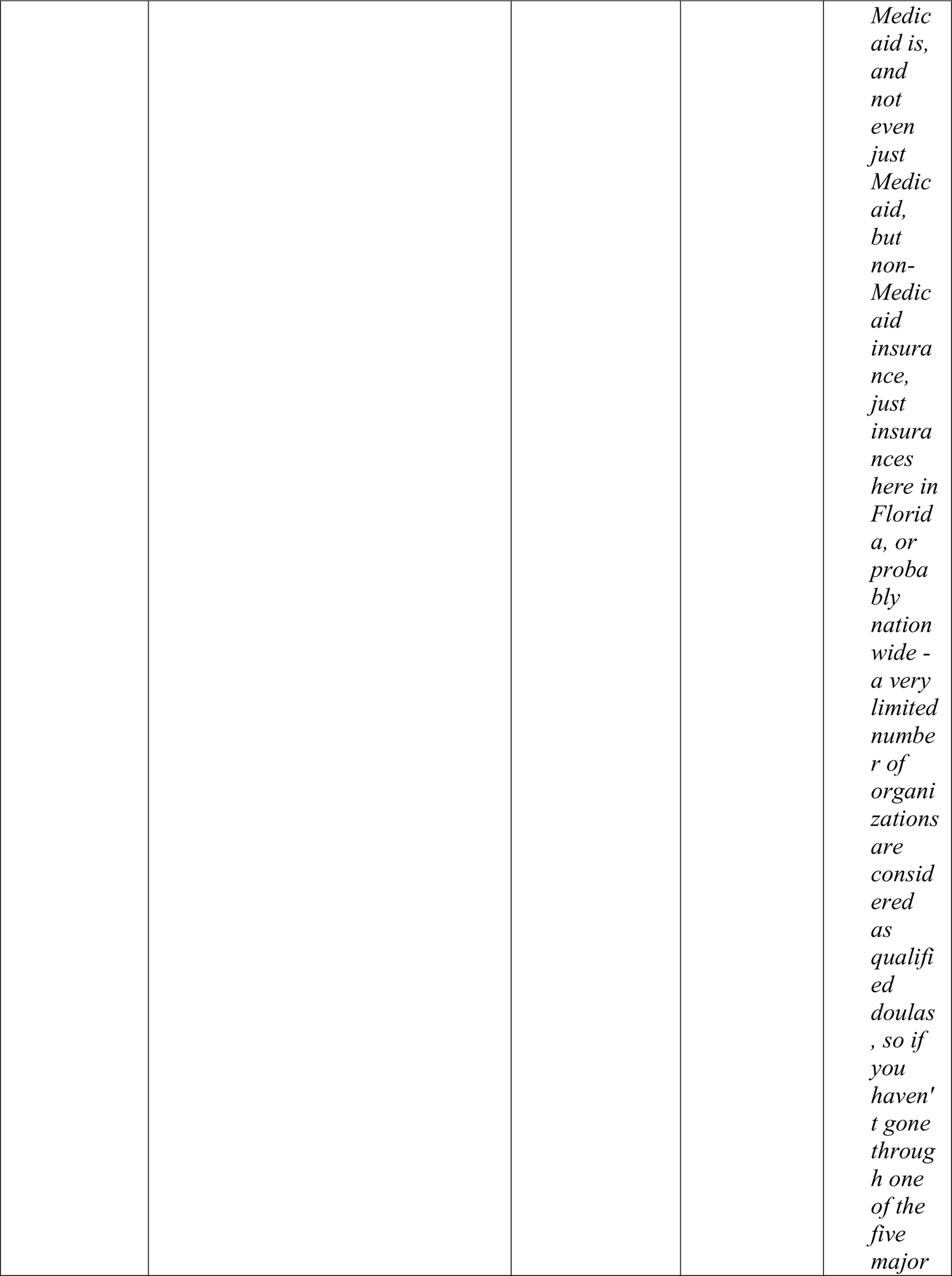
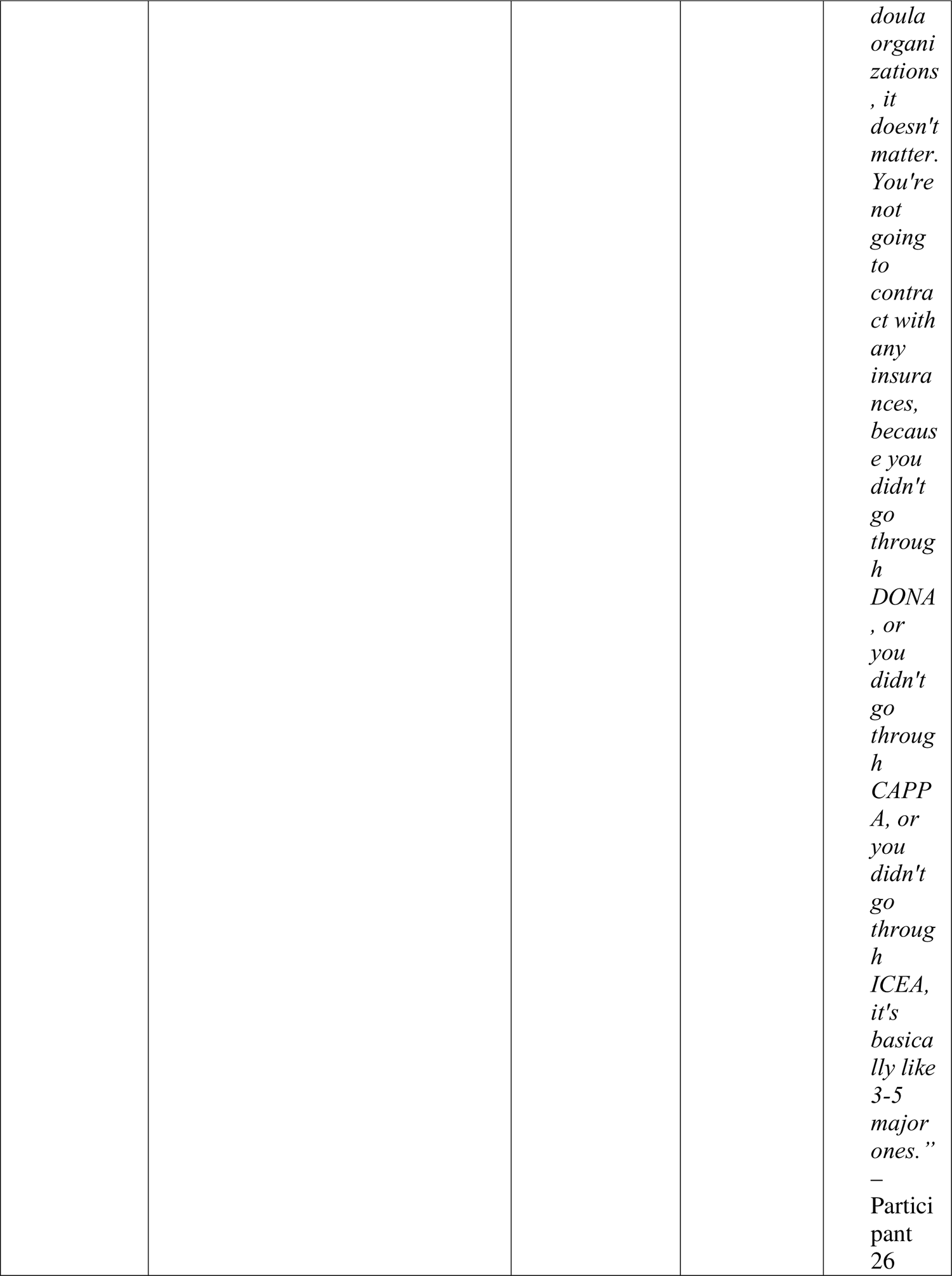
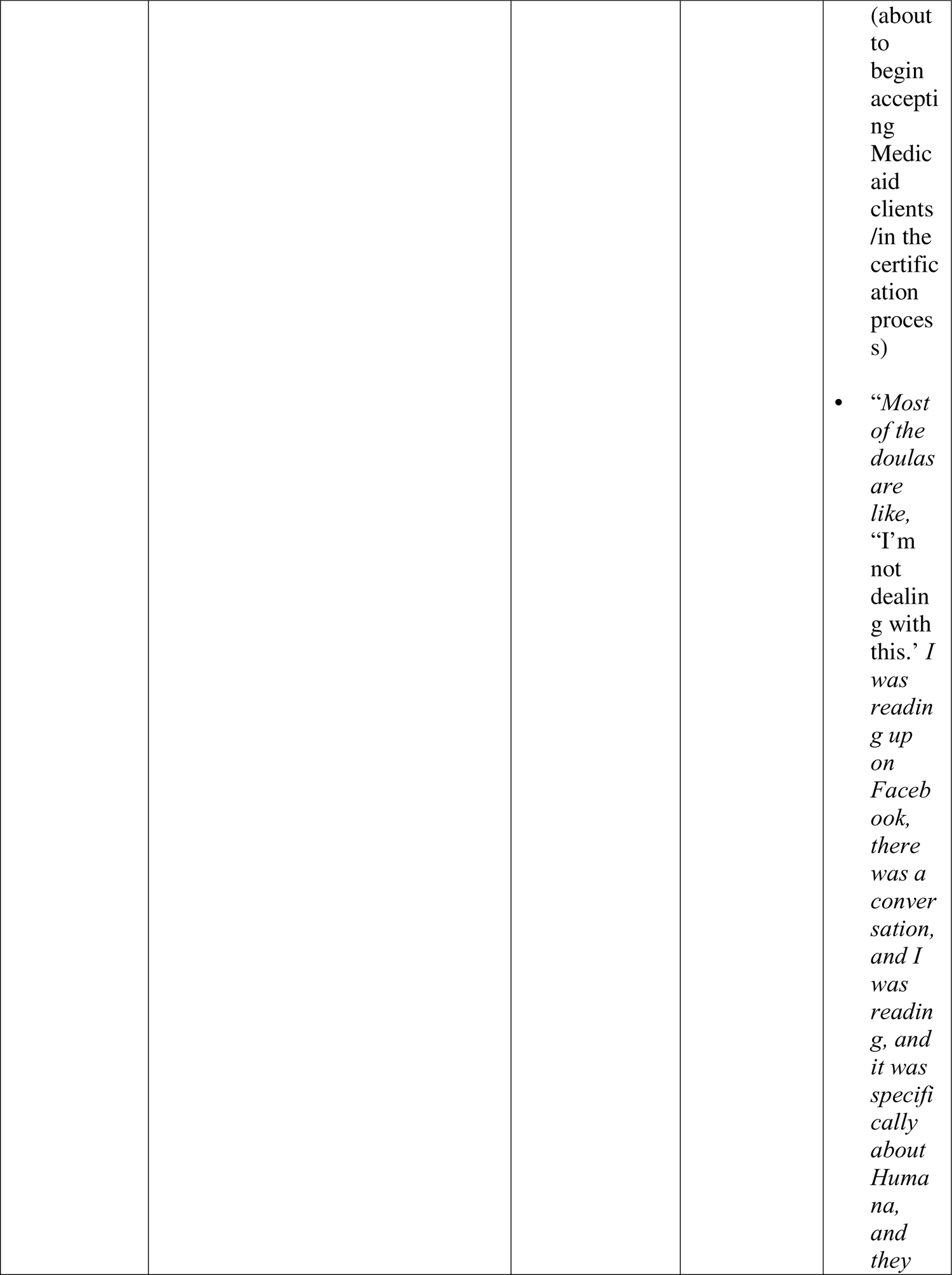
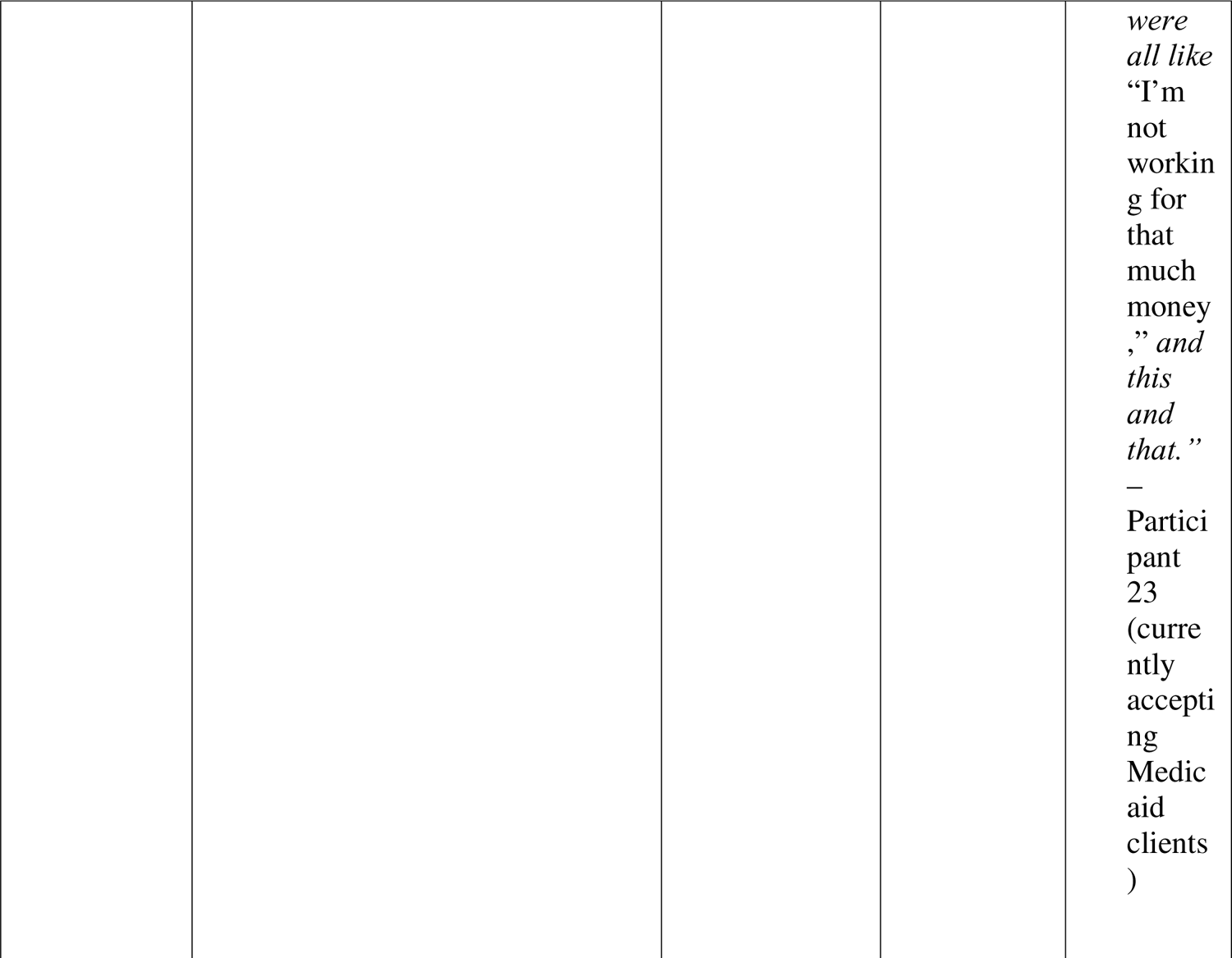
Perceived Barriers to Florida Medicaid Implementation Efforts. Includes applicable domains of the Theoretical Domains Framework (TDF), applicable TDF constructs, and supporting participant quotations.

**Table 3.** Perceived Facilitators to Florida Medicaid Implementation Efforts. Includes applicable domains of the Theoretical Domains Framework (TDF), applicable TDF constructs, and supporting participant quotations.

| **Perceived Facilitator (Theme)** | **Description of Facilitator** | **Domain(s) [definition(s)]** | **TDF Construct(s)** | **Illustrative Quotation(s)** |
| --- | --- | --- | --- | --- |
| Third parties help facilitate a smoother reimbursement process | third party structures (e.g., TDN) act as an intermediary to make the reimbursement process more navigable<br>TDN is the largest third party structure and most mentioned by participants | Environmental context and resources (any circumstance of a person’s situation or environment that discourages or encourages the | resources<br>facilitators | “*But the Doula Network handles all of it. My experience is very different than what XX* |
|  | • while TDN cannot make MCOs increase the speed at which they reimburse, they can act as an intermediary | development of skills and abilities, independence, social competence, and adaptive behavior) |  | *described as kind of like what people are saying, thankfully it's different. And so I was worried about all of those fears, that they'd be controlling, that I have to do it their way, all that. I have not found that to be true, I mean it's a little bit more tedious, it's a little bit more paperwork, it's a little bit more visits. However, I have found it extremely rewarding because the clients*<br>*that I get through [the Doula Network] - like I said, I've never had through private pay. So I feel like it's much more needed. Like when I'm at a birth for a client who is a Medicaid client or through the Doula Network I just feel like I'm doing the work, I'm doing the service, you know."* – Participant 30 (currently accepting Medicaid clients) |
| Aspirations for Medicaid accessibility/ dedication | doulas report seeing the Medicaid system as broken, but do not want to leave patients behind<br>feel that Medicaid in | Intentions (a conscious decision to perform a behavior or a resolve to act | stability of intentions<br>intrinsic motivation | "*The county that I live in, there's so many low-* |
| to clients | its current state is “smoke and mirrors”<br>• frustrations surrounding structure of the process, not the idea of Medicaid itself<br>• systemic breakdown of the Medicaid structure weighs heavily on doulas because they want to help their communities, but system makes it too difficult<br>• hence pro bono work, cash payments | in a certain way)<br>• Beliefs about capabilities (acceptance of the truth, reality, or validity about outcomes of a behavior in a given situation) | • self-efficacy<br>• beliefs | *income families who cannot afford support, so you know, being able to - most of them qualify for Medicaid, and so you know, being able to have the support that they need, that's really the only way. I also birthed my own babies using it [Medicaid], and I would not have been able to afford it and support myself and I know where I'm at personall*<br>*y, and that [a doula] would have been an amazing support system to have, and I know I did not consider it [a doula], and so I want to provide that for other women in my same position.”*<br>— Participant 6 (about to begin accepting Medicaid clients/in the certification process) |

### Barrier: reimbursement rates and doula livelihood

For all participants, the rates offered for doula services via Medicaid reimbursement posed significant obstacles to successful implementation of Medicaid services. All participants reflected on the low payouts offered by individual insurers/managed care organizations (MCOs). Frustration was reported surrounding the vast range in reimbursement rates ($300-$1,100, depending on the MCO’s negotiated rates) (Pasia, 2022). Several commented on the low payout rates not permitting for doulas to earn livable wages (*environmental context and resources*). For those currently working with Medicaid clients, comments centered on insufficient pay impeding doulas’ *own access* to proper healthcare, creating the need for establishing boundaries with the Medicaid system and their clients.

> “I gotta stop taking clients if you guys are not going to pay and I’m like, and I hate to do that, because I know these women have nobody and they need me – but at the same time, I have to pay rent.” (Participant 23)

In instances where participants had not worked with the Medicaid reimbursement process, they cited low payout rates and experiences they heard from other doulas as the reason they would not *directly* engage with Medicaid. Rather than utilizing the system, they reported getting more creative to avoid Medicaid while still helping the most in-need clients. Providing scaled pro bono services (while charging affluent clients more), accepting reduced rates via cash and PayPal, and having clients submit for their own reimbursement were commonly discussed as ways to avoid the hassle of Medicaid.

> “I don’t--I’m not on any insurance panels….and the problem is, it ranges so I can submit - I mean I always tell my clients if their insurance is accepted, I have my NPI and all that and I ask them what they need, I’ll give you the information, and you could submit it on your own. To me like, yes, I do some scaled or pro bono work per year but, overall, this is a business. I have two mouths to feed, not including my own, and I, at the end of the day, need to protect my family as well, so for me it’s not worth it to maybe get paid half of my fee.” (Participant 24)

### Barrier: reimbursement process

The reimbursement process was often labeled as a “nightmare” by those currently using it or those who had heard it discussed by other doulas experiencing it. One participant used to accept Medicaid clients but stopped due to her negative experience. The number one barrier to implementation regarding the Medicaid process was the time insurers take to reimburse doulas for their services. Participants reported waiting as long as two years to receive reimbursement after a client’s birth.

Participants also discussed the significant steps associated with filing for Medicaid reimbursement. The Medicaid process was mentioned as daunting for doulas as it requires them to navigate invoices, have charting capabilities, and develop a strong comprehension of multiple MCOs and their policies (*environmental context and resources*). MCOs may have codes for non-clinical support services (e.g., vaginal birth support, surgical birth support), but Health Care Financing Administration (HCFA) Current Procedural Terminology (CPT) codes do not exist specifically for doula reimbursement. Claims get rejected when incorrect information is entered, increasing frustrations. While third party networks exist to connect clients and doulas, and facilitate with filing doulas’ reimbursement paperwork, not all doulas utilize these services.

> “So it’s like another roadblock. They make it all so hard and complicated that all these people that we could be, you know, successfully working with them, helping them. But with these shitty rates and, like all this kind of process stuff we’re going through? Ugh. Just blah.” (Participant 1)

### Barrier: doula autonomy

Many participants reported that the individual requirements MCOs have put in place to 1) work with Medicaid clients and 2) receive reimbursement are at times not in alignment with their professional philosophies, meaning they fear their independence is being stripped away (*social/professional role and identity; social influences; emotion*). These comments often referenced doulas stating they did not want Medicaid telling them “how to do their jobs.”

> “I’ve heard it’s a nightmare [Medicaid], which is why I’ve not tried to look into it. I’ve heard that they pay what they want to pay when they want to pay. There’s a lot more expectation and a lot more. It’s just hard. It’s a lot more nerve-racking and you don’t have as much, like, freedom to practice, the way that you feel that you should be practicing or want to be practicing. And I just, I want to be able to be me and do what I do, how I do it, and I would rather, if someone came to me and was like, “oh hey, I have Medicaid.” I would rather take that client on as a no-pay versus whatever Medicaid is going to want to do.” (Participant 31)

### Barrier: certification criteria, or Medicaid requirements, viewed as unfair/unrealistic

Participants reported apprehensions surrounding certification criteria for working with Medicaid clients. No state or national certification criteria exist for doulas, but in Florida, doulas must be certified and credentialed to work with Medicaid clients. Each MCO determines which certifying organization they will contract with and accepts that organization’s criteria for certification. Doulas expressed frustration in being limited by the number of organizations that can provide certifications for working with Medicaid clients. Many doulas reported the initial steps required to become certified by MCOs (e.g., background screenings, liability insurance, letters of recommendation) as unfair and unrealistic (*reinforcement*).

> “Their expectations of what you have to have provided to be in network is ridiculous. You have to have a very specific Apple setup, you have to have very specific software…” (Participant 3)

### Barrier: misinformation

The Medicaid reimbursement process was described as a maze that doulas are trying to navigate. In attempting to describe the roadblocks in place for doulas, it became clear that a lack of direct, reputable information on how to effectively navigate the Medicaid process has created intense confusion and frustration for doulas (*knowledge*). Most participants in our sample reported not currently accepting Medicaid clients, citing their rationale as what they have heard about the process from other doulas. Upon further investigation in our stakeholder meetings, it became evident that the reimbursement process was so convoluted and ambiguous that doulas were relying *primarily* on anecdotal evidence about Medicaid because they had no other information source (*social influences*).

> “Most of the doulas are like “I’m not dealing with it.” I was reading up on Facebook, there was a conversation, and I was reading, and it was specifically about Humana, and they were all like “I’m not working for that much money,” and this and that. And I’m like, but I say it, but I’m thinking, but “somebody has to help them,” you know?” (Participant 23)

Doulas accepting Medicaid clients confirmed that Medicaid and individual MCOs did not, and have not, rolled out communication plans to ensure a more successful implementation for families, doulas, and clients. Thus, while there are truths embedded throughout a disorderly process, inaccuracies are being picked up, shared, and spread that may be causing further damage for the doula community’s buy-in of Medicaid reimbursement implementation.

Misinformation is defined as the unintentional sharing of inaccurate or misleading information (Suarez-Lledo & Alvarez-Galvez, 2021). Participants reported the doula community as being actively engaged with its peers, often referencing the cultivation of online support networks that are references as resources for their trusted information. Therefore, many participants reflected on anecdotal evidence about the Medicaid process that they had read in a Facebook group. Most often, doulas reported choosing not to accept Medicaid clients based on information they heard from other doulas. Many times, the information they reported was not accurate. A common example of misinformation provided by doulas included a claim that MCOs only work with the three to five largest certifying agencies. In reality, Florida has over 180 certifying organizations that MCOs may choose, but outdated information (based on processes from initial rollout in 2019) continues to be shared amongst doulas.

> “You have to do things essentially that are not within the scope of practice for doulas and they only are allowing very specific certifications for you to be in that network…” (Participant 3)

### Facilitator: structures to help facilitate billing and client engagement

For those participating in Medicaid reimbursement, all reflected on the necessity of having a third-party entity in place to help facilitate the process. Only one participant reported currently accepting Medicaid clients and having a positive experience with the reimbursement process. TDN was referenced as a popular entity used by participants to mitigate the frustrations surrounding paperwork. While these third parties cannot increase the speed at which insurers reimburse, they can oversee the filing for a complex system (*environmental context and resources*).

### Facilitator: (cautious) aspirations for Medicaid accessibility

All participants, regardless of their experiences with or perspectives on the Medicaid system in relation to doula care, have an ardent desire to make Medicaid truly accessible for all birthing families. Comments centered on Medicaid being *available*, but not *accessible*, presenting a “smoke and mirrors” scenario where the state, and insurers, can claim they are helping families, but the execution of this initiative needs significant enhancement (*beliefs about capabilities*).

> “It’s a broken system. And it’s a system that is not favorable to all types of care, and in my opinion, the care that is necessary for moms to have, for families to have good experiences and a good start to their new family. So yeah, it’s very unfortunate. I was one of those Medicaid moms when I had my own daughter, and I was thankful enough to find a midwifery practice that would accept what I had. But it took some searching to find what I needed. So it is difficult.” (Participant 29)

## Discussion and Conclusion

Low reimbursement rates prohibiting sustainable livelihoods, an unnavigable reimbursement process, concerns for professional autonomy, certification and Medicaid requirements perceived as unrealistic, and significant misinformation were identified as barriers to doulas’ satisfaction with and subsequent uptake of Medicaid services. Identified facilitators included third party assistance to supplement billing and client engagement, as well as doulas’ desires to help community members who are most in need.

The TDF illuminated an intersection of multiple factors that may be contributing to doulas’ action or inaction regarding the implementation of Medicaid reimbursement for doula services. *Knowledge* is the first (and most critical) entry point for necessary intervention, as doulas need more information on the reimbursement process, whether currently accepting Medicaid clients or wanting an entry point to the process. One key example of misinformation centered on certifications. Many justified their decision to not work with Medicaid because of an assumption that they could only receive a certification from three to five of the largest certifying agencies (e.g., DONA, CAPPA, ICEA). Stakeholder team meetings revealed that this information *was true at one time*, when doula services began to be covered by Medicaid in 2019. While MCOs *can* dictate the certifying organization, we found that misinformation was occasionally spread amongst our doula sample, but not with ill intent. MCOs often release little, if any, new information to doulas to help clarify the process or make it more navigable. Thus, doulas currently do not have one trusted, reliable source for Medicaid reimbursement information that could help to refute untrue claims or educate a new workforce.

Findings from this study offer insight into doula perceptions of Medicaid in Florida and beyond. While information sources are limited for those navigating this space, our data revealed a series of applied considerations for doulas wanting to engage with Medicaid reimbursement (provided as a supplementary table). Across the United States, doula services are being examined as an added benefit option to support populations made vulnerable. The findings from this study will not only continue to influence the development of this benefit in Florida, but nationwide as well. As of 2023, 14 states are currently reimbursing for doula services, 11 are in process or advocating for legislative action surrounding doula services, and 13 states are actively convening stakeholders to improve or develop recommendations for implementing doula coverage (Chen, 2022; Robles-Fradet, 2021). Our findings can play a role in shaping policy in the areas of stakeholder engagement, billing processes and procedures, certification or credentialing requirements, and resources needed for doula support. As leading doula certifying organizations continue to expand their membership across the world (Muza, 2018), and doulas serve to help mothers navigate language, cultural, and healthcare barriers, the global implications of this work are important when considering how implementation of services can best be refined for doulas to help more vulnerable families.

The literature provides evidence on how doulas positively influence pregnancy (McLeish & Redshaw, 2019), birth (Bohren et al., 2017; K. J. Gruber et al., 2013), and the postpartum period, (Bohren et al., 2017; McLeish & Redshaw, 2019) and how they can particularly impact outcomes for marginalized birthing people (Falconi et al., 2022). As national MM rates rise, and the United States seeks answers to respond to the maternal health crisis, it is critical that the systems supporting our most vulnerable populations are optimized. The Medicaid expanded benefit to include doula care provides critical support to those most at risk for MM, SMM, and infant mortality; however, reimbursement rates need to ensure a livable wage for doulas and the billing process should be optimized for coverage to be truly accessible. Most importantly, and consistent with the reproductive justice framework, doulas can help advocate for pregnant people who consistently face oppression in the healthcare system, so that their needs and voices are heard in spaces where they have been discounted. Data-driven policies must be utilized for the development and implementation of doula benefits to ensure they are impactful for those facing oppression in healthcare. To be able to provide such services, it is critically important that primary information sources containing clear billing processes and consistent coding be made available for doulas.

## Data Availability

All data produced in the present study upon reasonable request to the authors.

## Acknowledgements

We thank Elizabeth Simmons and Sharon Lonix from The Doula Network for their expertise and support of this work.

