## Supplemental Table. for "Applying the Theoretical Domains Framework to the Implementation of Medicaid Coverage for Doula Services: Doulas’ Perceptions of Barriers and Facilitators in the State of Florida"

**Supplementary Table.**
Considerations for Doulas Navigating Florida’s Medicaid Process

| **Consideration** | **Description** |
| --- | --- |
| 1. Obtain a National Provider Identifier (NPI) number | - a doula can obtain this number on their own - “doula” is a taxonomy code option |
| 2. Obtain a Medicaid ID number | - obtaining this makes a doula legally capable of receiving reimbursement - In Florida, a MCO has to apply for this on behalf of a doula, meaning that a doula *does not have* an avenue to apply for this independently (must already be contracted with a group) - getting a Medicaid ID number does not guarantee you will get paid |
| 3. Become educated on how to get MCO/insurance contracts | - take the path a provider would to become credentialed, but take the route as a doula:   - visit MCO websites and engage with their provider resources |
| 4. Become educated on billing | - if not utilizing a third party (like The Doula Network), you will depend on the programming and resources of the MCO because how you will for one (e.g., Humana) will not be how you bill for another (e.g., BCBS)   - however, this information is not always readily available or easily accessible (why it would be ideal for MCOs to have “doula pages” with information available on their sites) |
| 5. Become educated on codes | - there is no code for doula services, but there are codes for what is happening - become educated on the group of codes you have to bill for (e.g., vaginal birth) |
| 6. Understand state requirements for being a Medicaid provider | - this area is often overlooked, but very important - Medicaid has several requirements that doulas are responsible for following. - requirements regarding record storage (paper vs. electronic), length of record storage. Doulas must learn about this process to ensure compliance. The Centers for Medicare & Medicaid (CMS) can review several years after and revoke payment if requirements are not followed, meaning doulas need to be able to navigate this terrain. - charting (even if working with a third party to help facilitate) is another state requirement (understanding the regulations surrounding privacy and documentation associated with taking federal money) |
